# Balanced chromosomal rearrangements offer insights into coding and noncoding genomic features associated with developmental disorders

**DOI:** 10.1101/2022.02.15.22270795

**Authors:** Chelsea Lowther, Mana M. Mehrjouy, Ryan L. Collins, Mads C. Bak, Olga Dudchenko, Harrison Brand, Zirui Dong, Malene B. Rasmussen, Huiya Gu, David Weisz, Lusine Nazaryan-Petersen, Amanda S. Fjorder, Yuan Mang, Allan Lind-Thomsen, Juan M.M. Mendez, Xabier Calle, Anuja Chopra, Claus Hansen, Merete Bugge, Roeland V. Broekema, Teppo Varilo, Tiia Luukkonen, John Engelen, Angela M. Vianna-Morgante, Ana Carolina S. Fonseca, Juliana F. Mazzeu, Halinna Dornelles-Wawruk, Kikue T. Abe, Joris R. Vermeesch, Kris Van Den Bogaert, Carolina Sismani, Constantia Aristidou, Paola Evangelidou, Albert A. Schinzel, Damien Sanlaville, Caroline Schluth-Bolard, Vera M. Kalscheuer, Maren Wenzel, Hyung-Goo Kim, Katrin Õunap, Laura Roht, Susanna Midyan, Maria C. Bonaglia, Anna Lindstrand, Jesper Eisfeldt, Jesper Ottosson, Daniel Nilsson, Maria Pettersson, Elenice F. Bastos, Evica Rajcan-Separovic, Fatma Silan, Frenny J. Sheth, Antonio Novelli, Eirik Frengen, Madeleine Fannemel, Petter Strømme, Nadja Kokalj Vokač, Cornelia Daumer-Haas, Danilo Moretti-Ferreira, Deise Helena de Souza, Maria A. Ramos-Arroyo, Maria M. Igoa, Lyudmila Angelova, Peter M. Kroisel, Graciela del Rey, Társis A.P. Vieira, Suzanne Lewis, Wang Hao, Jana Drabova, Marketa Havlovicova, Miroslava Hancarova, Zdeněk Sedláček, Ida Vogel, Tina D. Hjortshøj, Rikke S. Møller, Zeynep Tümer, Christina Fagerberg, Lilian B. Ousager, Bitten Schönewolf-Greulich, Mathilde Lauridsen, Juliette Piard, Celine Pebrel-Richard, Sylvie Jaillard, Nadja Ehmke, Eunice G. Stefanou, Czakó Marta, Kosztolányi György, Ashwin Dalal, Usha R. Dutta, Rashmi Shukla, Fortunato Lonardo, Orsetta Zuffardi, Gunnar Houge, Doriana Misceo, Shahid M. Baig, Alina Midro, Natalia Wawrusiewicz-Kurylonek, Isabel M. Carreira, Joana B. Melo, Laura Rodriguez Martinez, Miriam Guitart, Lovisa Lovmar, Jacob Gullander, Kerstin B.M. Hansson, Cynthia de Almeida Esteves, Yassmine Akkari, Jacqueline R. Batanian, Xu Li, James Lespinasse, Asli Silahtaroglu, Christina Halgren Harding, Lotte Nylandsted Krogh, Juliet Taylor, Klaus Lehnert, Rosamund Hill, Russell G. Snell, Christopher A. Samson, Jessie C. Jacobsen, Brynn Levy, Ozden Altiok Clark, Asli Toylu, Banu Nur, Ercan Mihci, Kathryn O’Keefe, Kiana Mohajeri-Stickels, Ellen S. Wilch, Tammy Kammin, Raul E. Piña-Aguilar, Katarena Nalbandian, Sehime G. Temel, Sebnem Ozemri Sag, Burcu Turkgenc, Arveen Kamath, Adriana Ruiz-Herrera, Siddharth Banka, Samantha L.P. Schilit, Benjamin B. Currall, Naomi Yachelevich, Stephanie Galloway, Wendy K. Chung, Salmo Raskin, Idit Maya, Naama Orenstein, Nesia Kropach Gilad, Kayla R. Flamenbaum, Beverly N. Hay, Cynthia C. Morton, Eric Liao, Kwong Wai Choy, James F. Gusella, Peter Jacky, Erez Lieberman Aiden, International Breakpoint Mapping Consortium (IBMC), Danish Cytogenetic Central Registry Study Group, Developmental Genome Anatomy Project (DGAP), Iben Bache, Michael E. Talkowski, Niels Tommerup

**Affiliations:** Center for Genomic Medicine, Massachusetts General Hospital, Boston, MA, USA; Program in Medical and Population Genetics, Broad Institute of MIT and Harvard, Boston, MA, USA; Department of Neurology, Harvard Medical School, Boston, MA, USA; Department of Cellular and Molecular Medicine, University of Copenhagen, Copenhagen, Denmark; Program in Bioinformatics and Integrative Genomics, Division of Medical Sciences, Harvard Medical School, Boston, MA, USA; The Center for Genome Architecture, Department of Molecular and Human Genetics, Baylor College of Medicine, Houston, TX 77030, USA; Center for Theoretical Biological Physics and Department of Computer Science, Rice University, Houston, TX 77030, USA; Department of Obstetrics and Gynaecology, The Chinese University of Hong Kong, Hong Kong, China; Department of Clinical Genetics, Copenhagen University Hospital - Rigshospitalet, Copenhagen, Denmark; Institute of Biological Psychiatry, Mental Health Services, Copenhagen University Hospital, Copenhagen, Denmark; Precision Diagnostics, Ahmedabad, India; Department of Biology and Medical Genetics, Charles University Second Faculty of Medicine and University Hospital Motol, Prague, Czech Republic; Department of Medical Genetics, University of Helsinki, Helsinki, Finland; Population Health Unit, Department of Public Health and Welfare, National Institute for Health and Welfare, Helsinki, Finland; Institute for Molecular Medicine Finland (FIMM), Biomedicum 2U, Helsinki, Finland; Maastricht University Medical Center, Maastricht, Netherlands; Departamento de Genética e Biologia Evolutiva, Instituto de Biociências, Universidade de São Paulo, São Paulo, Brazil; Faculdade de Medicina, Universidade de Brasília, Brasília, Brazil; Cytogenetic Laboratory Molecular Pathology, SARAH Network of Rehabilitation Hospitals, Brasília, Brazil; Center for Human Genetics, KU Leuven, Leuven, Belgium; Department of Cytogenetics and Genomics, The Cyprus Institute of Neurology and Genetics, Nicosia, Cyprus; The Cyprus School of Molecular Medicine, The Cyprus Institute of Neurology and Genetics, Nicosia, Cyprus; Institute of Medical Genetics, University of Zurich, Schlieren, Switzerland; Service de Génétique, Hospices Civils de Lyon, Groupement Hospitalier Universitaire, Bron, France; Institut NeuroMyoGene, Université Claude Bernard Lyon, Bron, France; Group Development and Disease, Max Planck Institute for Molecular Genetics, Berlin, Germany; Neurological Disorders Research Center, Qatar Biomedical Research Institute, Hamad Bin Khalifa University, Doha, Qatar; Department of Clinical Genetics, United Laboratories Tartu University Hospital, Tartu, Estonia; Institute of Clinical Medicine, University of Tartu, Tartu, Estonia; Center of Medical Genetics and Primary Health Care, Yerevan, Armenia; Cytogenetics Laboratory, Scientific Institute, IRCCS Eugenio Medea, Bosisio Parini, Lecco, Italy; Department of Molecular Medicine and Surgery, Center for Molecular Medicine, Karolinska Institutet, Stockholm, Sweden; Department of Clinical Genetics, Karolinska University Hospital, Stockholm, Sweden; Science for Life Laboratory, Karolinska Institutet Science Park, Solna, Sweden; Clinical Genetics and Genomics, Sahlgrenska University Hospital, Gothenburg, Sweden; Laboratorio de Citogenética Clínica - Centro de Genética Médica, Instituto Nacional Fernandes Figueira - Fiocruz, Rio de Janeiro, Brazil; Department of Pathology and Laboratory Medicine, University of British Columbia, Vancouver, BC Canada; Department of Medical Genetics, Canakkale 18 March University Medical Faculty, Canakkale 18 March University, Research Hospital, Medical Genetic Lab, Canakkale, Turkey; FRIGE Institute of Human Genetics, Ahmedabad, India; Translational Cytogenomics Research Unit, Bambino Gesù Children’s Hospital, IRCCS, Rome, Italy; Department of Medical Genetics, Oslo University Hospital and University of Oslo, Olso, Norway; Division of Pediatric and Adolescent Medicine, Oslo University Hospital and University of Oslo, Olso, Norway; Laboratory of Medical Genetics, University Medical Centre Maribor, Maribor, Slovenia; Medical Faculty University of Maribor, Maribor, Slovenia; Prenatal-Medicine Munich, Friedenheimer Brücke, München, Germany; Chemical and Biological Sciences Department, São Paulo State University (UNESP), São Paulo, Brazil; Servicio de Genética, Hospital Universitario de Navarra, SNS-Osasunbidea, C/ Irunlarrea 4, Pamplona, Navarra, Spain; Laboratory of Medical Genetics, St Marina University Hospital, Varna, Bulgaria; Department of Medical Genetics, Medical University of Varna, Bulgaria; Diagnostic and Research Institute for Human Genetics, Medical University of Graz, Graz, Austria; Centro de Investigaciones Endocrinológicas “Dr César Bergadá”, CONICET. FEI, Hospital de Niños “Ricardo Gutiérrez”, Buenos Aires, Argentina; Department of Medical Genetics, State University of Campinas (Unicamp), Campinas, São Paulo, Brazil; Department of Medical Genetics, The University of British Columbia, Vancouver, BC, Canada; Department of Cell Biology and Medical Genetics, School of Medicine, Zhejiang University, Zhejiang, China; Prenatal Diagnosis Center, Hangzhou Women’s Hospital, Hangzhou, China; Department of Clinical Genetics, Aarhus University Hospital, Aarhus, Denmark; Department of Clinical Medicine, Aarhus University, Aarhus, Denmark; Department of Epilepsy Genetics and Personalized Medicine, Danish Epilepsy Centre, Filadelfia, Dianalund and University of Southern Denmark, Odense, Denmark; Kennedy Center, Department of Clinical Genetics, Copenhagen University Hospital, Copenhagen, Denmark; Department of Clinical Medicine, Faculty of Health and Medical Sciences, University of Copenhagen, Denmark; Department of Clinical Genetics, Odense University Hospital, Odense, Denmark; Department of Clinical Research, University of Southern Denmark, Odense, Denmark; Department of Clinical Genetics, Vejle Hospital, Copenhagen, Denmark; Centre de Génétique Humaine, Centre Hospitalier Régional Universitaire, Université de Franche-Comté, Besançon, France; CHRU Clermont-Ferrand, Clermont, France; Service de Cytogénétique et Biologie Cellulaire, CHU Rennes, Rennes, France; Institute of Medical Genetics and Human Genetics, Augustenburger Platz, Berlin, Germany; Lab of Medical Genetics, University General Hospital of Patras, Patras, Greece; Department of Medical Genetics, University of Pécs Medical School, Pecs, Hungary; Diagnostics Division, Centre for DNA Fingerprinting and Diagnostics, Hyderabad, India; Department of Pediatrics, Division of Genetics, All India Institute of Medical Sciences, New Delhi, India; Medical Genetics Unit, A.O.R.N. “San Pio” - P.O. “G. Rummo”, Benevento, Italy; Department of Molecular Medicine, University of Pavia, Pavia, Italy; Department of Medical Genetics, Haukeland University Hospital, Bergen, Norway; Human Molecular Genetics Laboratory, National Institute for Biotechnology and Genetic Engineering (NIBGE), Faisalabad, Pakistan; Department of Clinical Genetics, Medical University of Bialystok, Bialystok, Poland; Cytogenetics and Genomics Laboratory, Clinical Academic Center of Coimbra, iCBR/CIMAGO-CIBB, Faculty of Medicine, University of Coimbra, Coimbra, Portugal; Technical Director of the Laboratory of Genetics and Molecular Biology, AbaCid laboratory, Madrid, Spain; HM Hospitales, Madrid, Spain; Genetics Laboratory, UDIAT-Centre Diagnòstic, Parc Taulí Hospital Universitari, Institut d’Investigacioó i Innovació Parc Taulí I3PT, Universitat Autònoma de Barcelona, Sabadell, Spain; Department of Clinical Genetics, Skåne Regional and University Laboratories Hospital, Lund, Sweden; Department Clinical Genetics, Leiden University Medical Center, Leiden, Netherlands; Militar Hospital, Montevideo, Ururguay; Cytogenetics and Molecular Pathology, Legacy Laboratory Services, Portland, OR, USA; Department of Pediatrics and Pathology, Saint Louis University Medical Center, Saint Louis, MO, USA; TPMG Regional Genetics Laboratory, Kaiser Permanente, San Jose, CA, USA; Service de Genetique, Centre Hospitalier Metropole Savoie, Chambery, France; Wilhelm Johannsen Centre for Functional Genome Research, Department of Cellular and Molecular Medicine, University of Copenhagen, Copenhagen, Denmark; Genetic Health Service New Zealand, Auckland City Hospital, Auckland, New Zealand; Centre for Brain Research and School of Biological Sciences, The University of Auckland, Auckland, New Zealand; Department of Neurology, Auckland City Hospital, Auckland, New Zealand; Department of Pathology and Cell Biology, Columbia University Medical Center, New York City, NY, USA; Department of Medical Genetics, Faculty of Medicine, Akdeniz University, Antalya, Turkey; Department of Pediatrics, Division of Pediatric Genetics, Faculty of Medicine, Akdeniz University, Antalya, Turkey; Department of Obstetrics and Gynecology, Brigham and Women’s Hospital, Boston, MA, USA; Department of Pathology, Brigham and Women’s Hospital, Boston, MA, USA; PhD Program in Biological and Biomedical Sciences, Harvard Medical School, Boston, MA, USA; Instituto de Ciencias en Reproducción Humana, León, México; Massachusetts College of Pharmacy and Health Sciences, Boston, MA, USA; Department of Medical Genetics, Bursa Uludag University, Bursa, Turkey; The All Wales Medical Genomics Service (AWMGS), Institute of Medical Genetics, University Hospital of Wales, Cardiff, Wales; Department of Medical Genetics, Hospital de Especialidades Pediátrico León, Guanajuato, México; Division of Evolution, Infection and Genomics, School of Biological Sciences, Faculty of Biology Medicine and Health, University of Manchester, Manchester, UK; Manchester Centre for Genomic Medicine, St Mary’s Hospital, Manchester University NHS Foundation Trust, Health Innovation Manchester, Manchester, UK; Laboratory for Molecular Medicine, Mass General Brigham Personalized Medicine, Cambridge, MA, USA; Clinical Genetics Services, Department of Pediatrics, New York University School of Medicine, New York, NY, USA; Department of Maternal Fetal Medicine, Women’s Genetics, Columbia University, New York City, NY, USA; Departments of Pediatrics and Medicine, Columbia University, New York City, NY, USA; Laboratorio Genetika, Curitiba, Parana, Brazil; Recanati Genetic Institute, Rabin Medical Center, Beilinson Hospital, Petah Tikva, Israel; Sackler Faculty of Medicine, Tel Aviv University, Tel Aviv, Israel; Pediatric Genetic Unit, Schneider Children’s Medical Center of Israel, Petah Tikvah, Israel; University of Toronto, The Prenatal Diagnosis and Medical Genetics Program, Department of Obstetrics and Gynecology, Mount Sinai Hospital, University of Toronto, Toronto, Ontario, Canada; Division of Genetics, Department of Pediatrics, UMass Chan Medical School, UMass Memorial Health, Worcester MA; Division of Plastic and Reconstructive Surgery, Mass General Brigham, Harvard Medical School, Boston, MA, USA; Shriners Hospital for Children, Boston, MA, USA; Molecular Neurogenetics Unit, Center for Genomic Medicine, Massachusetts General Hospital, Boston, MA, USA; NW Permanente, PC, Emeritus, Portland, OR, USA; UWA School of Agriculture and Environment, The University of Western Australia, Crawley, WA 6009, Australia; Broad Institute of MIT and Harvard, Cambridge, MA 02139, USA; Shanghai Institute for Advanced Immunochemical Studies, ShanghaiTech, Pudong 201210, China; Division of Molecular Genetics, ICMS, Fujita Health University, Aichi, Japan; Genetics Department, Centre Hospitalier Universitaire, Angers, France; Department of Genetics, Alexandra General Hospital, Athens, Greece; Medical School, National an Kapodistrian University of Athens, Athens, Greece; National Institute of Mental Health and Neurosciences, Bangalore, India; Medical Genetics, Institute of Medical Genetics and Pathology, University Hospital Basel and University of Basel, Basel, Switzerland; Human Cytogenetics Department, National Research Centre, Cairo, Egypt; Dept Pathology and Laboratory Medicine, Medical University of South Carolina, Charleston, SC, USA; Cytogenetics, Dammam Regional Lab, Dammam, Saudi Arabia; Laboratório de Citogenética, Humana e Genética Molecular, PUC Goiás, LACEN/SES-GO, Goiaani, Brazil; CSIR-Centre for Cellular and Molecular Biology, Hyderabad, India; Istanbul Medical Faculty Medical Genetics Department, Istanbul, Turkey; Department of Medical Biology and Genetics, Faculty of Medicine, Dokuz Eylul University, Izmir, Turkey; Department of Medical Genetics, School of Medicine, Kayseri, Turkey; Human Genome Center, School of Medical Sciences, Health; Campus, Universiti Sains Malaysia, Kubang Kerian, Kelantan, Malaysia; UKM Medical Molecular Biology Institute, Universiti Kebangsaan Malaysia, Kuala Lumpur, Malaysia; Division of Medical Genetics, Department of Pediatrics, Hospital Santa Maria, Centro Hospitalar Universitário Lisboa Norte, Lisbon, Portugal; Department of Basic Immunology, Faculty of Medicine, Universidade de Lisboa, Lisbon, Portugal; Department of Medical Genetics, Centro Hospitalar Universitário Lisboa Norte, Lisbon, Portugal; School of Medicine and Surgery, University Milan-Bicocca, Milan, Italy; Laboratory of Medical Cytogenetics and Molecular Genetics, IRCCS Istituto Auxologico Italiano, Milan, Italy; Unit of Chromosomal Genetics and Research Plateform Chromostem, Arnaud de Villeneuve Hospital, Montpellier, France; Laboratory of Medical Genetics, San Gerardo Hospital, Monza, Italy; Cytogenetics laboratory, CBGC, Hospital C.U. Virgen de la Arrixaca, Murcia, Spain; Department of Cytogenetics, APHP centre-Université de Paris, Hopital Cochin, Paris, France; Cytogenetics Department, Genomedica, Piraeus, Greece; Cytogenomic Laboratory, Department of Pathology, Faculdade de Medicina FMUSP, Universidade de Sao Paulo, Sao Paulo, Brazil; Genetics Division, Universidade Federal de São Paulo, São Paulo, Brazil; Center for Biomedical Research, Faculty of Medicine, Diponegoro University, Semarang, Indonesia; Research Centre for Genetic Engineering and Biotechnology “Georgi D. Efremov”, Macedonian Academy of Sciences and Arts, Skopje, Republic of North Macedonia; Nationa Genetic Laboratory, Medical University Sofia, UHOG “Maichin dom”, Sofia, Bulgaria; Department of Biology, Medical Genetics and Microbiology, Faculty of Medicine, Sofia University St. Kliment Ohridski, Sofia, Bulgaria; University Hospital “Lozenets”, Dean’s building, Laboratory of Medical Genetics and Molecular Biology, Sofia, Bulgaria; Department of Pathology, Faculty of Medicine, Prince of Songkla University, Songkhla, Thailand; Laboratoire de Génétique médicale, Université de Lorraine, Vandoeuvre-les-Nancy, France; Department of Human and Medical Genetics, Institute of Biomedical Sciences, Faculty of Medicine, Vilnius University, Vilnius, Lithuania; Centre of Excellence for Reproductive and Regenerative Medicine, Children’s Hospital Zagreb, Medical School University of Zagreb, Zagreb, Croatia; Department of Assisted Reproduction and Genetics, Jaslok-FertilTree International Fertility Centre, Jaslok Hospital and Research Centre, Mumbai, India; Department of Genetics, Blavatnik Institute, Harvard Medical School, Boston, MA, USA; Department of Reproductive Biology and Stem Cells, Institute of Human Genetics, Polish Academy of Sciences, Poznan, Poland; Department of Medical Biology and Genetics, Faculty of Medicine, Istanbul Aydin University, Istanbul, Turkey; Genetikum, Genetic Counseling and Diagnostics, Genetikum Neu-Ulm, Neu-Ulm, Germany

## Abstract

Balanced chromosomal rearrangements (BCRs), including inversions, translocations, and insertions, reorganize large sections of the genome and contribute substantial risk for developmental disorders (DDs). However, the rarity and lack of systematic screening for BCRs in the population has precluded unbiased analyses of the genomic features and mechanisms associated with risk for DDs versus normal developmental outcomes. Here, we sequenced and analyzed 1,420 BCR breakpoints across 710 individuals, including 406 DD cases and the first large-scale collection of 304 control BCR carriers. We found that BCRs were not more likely to disrupt genes in DD cases than controls, but were seven-fold more likely to disrupt genes associated with dominant DDs (21.3% of cases vs. 3.4% of controls; *P =* 1.60×10^−12^). Moreover, BCRs that did not disrupt a known DD gene were significantly enriched for breakpoints that altered topologically associated domains (TADs) containing dominant DD genes in cases compared to controls (odds ratio [OR] = 1.43, *P =* 0.036). We discovered six TADs enriched for noncoding BCRs (false discovery rate < 0.1) that contained known DD genes (*MEF2C, FOXG1, SOX9, BCL11A, BCL11B*, and *SATB2*) and represent candidate pathogenic long-range positional effect (LRPE) loci. These six TADs were collectively disrupted in 7.4% of the DD cohort. Phased Hi-C analyses of five cases with noncoding BCR breakpoints localized to one of these putative LRPEs, the 5q14.3 TAD encompassing *MEF2C*, confirmed extensive disruption to local 3D chromatin structures and reduced frequency of contact between the *MEF2C* promoter and annotated enhancers. We further identified six genomic features enriched in TADs preferentially disrupted by noncoding BCRs in DD cases versus controls and used these features to build a model to predict TADs at risk for LRPEs across the genome. These results emphasize the potential impact of noncoding structural variants to cause LRPEs in unsolved DD cases, as well as the complex interaction of features associated with predicting three-dimensional chromatin structures intolerant to disruption.

## INTRODUCTION

Balanced chromosomal rearrangements (BCRs), including translocations, insertions, and inversions, are a unique class of rare genomic variation that occur roughly five-fold more frequently in individuals with developmental disorders (DDs) than in the general population.^1–6^ Delineation of BCR breakpoints has long represented an approach to discover novel disease genes,^7,8^ and has been accelerated by innovative methods using whole genome sequencing (WGS) with long-inserts (liWGS) to capture BCR breakpoints.^9–16^ Our previous WGS analyses suggested that 26.6% of cytogenetically visible BCRs contribute to risk for DDs due to direct gene disruption.^11^ The observation, while substantial, also implies that alternative mechanisms of disease are likely to be mediating additional genetic risk for DDs due to BCRs. However, the lack of sufficient sample sizes, and the virtual absence of large cohorts of unaffected control BCR carriers with sequence-resolved breakpoints, have precluded a systematic evaluation of the full spectrum of pathogenic mechanisms associated with BCRs to date.

Beyond the direct disruption of disease-associated protein-coding genes, emerging evidence has begun to implicate a small number of noncoding elements in the etiology of DDs. Recent analyses have emphasized the challenges with the statistically rigorous genome-wide discovery of rare and *de novo* noncoding regulatory risk variants, including their small average effect sizes, the lack of a cipher equivalent to trinucleotide codons for variant interpretation, and the large number of noncoding functional categories that could be tested.^17–19^ Nonetheless, there are examples of noncoding variants with strong regulatory consequences and considerable influence on risk for DDs,^19–21^ including long-range positional effects (LRPEs) that result from disruption of or topological associating domains (TADs)^20,24–26^ or long intergenic noncoding RNAs (lincRNAs).^22,23^ The disruption of TADs, which are megabase-sized regulatory domains of folded chromatin that contain most *cis*-regulatory interactions,^27–30^ can lead to loss of physical connections between enhancers and their target genes^24,25^ and/or the generation of ectopic enhancer-promoter contacts through a process known as “enhancer adoption”.^31,32^ While relatively few studies have systematically evaluated the contribution of rare noncoding variants to risk for disease, BCRs represent a unique class of highly penetrant genomic variation from which we might begin to understand the mechanisms of pathogenic noncoding variants in DDs given the outsized impact of BCRs on genome structure and function.^11,32,33^

In this study, we analyzed 1,420 BCR breakpoints from 710 unrelated individuals, including 406 DD cases as well as the first large-scale sequence-resolved cohort of 304 unaffected BCR carriers (*i*.*e*., controls), and evaluated a range of mechanisms by which BCRs may increase risk for DDs. Our analyses revealed a series of significant chromosomal, genic, and noncoding loci associated with DDs, as well as features that distinguished BCRs occurring in DD cases versus unaffected controls. In addition to refining DD risk estimates for BCRs directly disrupting dominant DD genes, we identified six TADs significantly associated with recurrent disruption by noncoding BCRs in DD cases, representing strong-effect LRPEs in DDs. Collectively, 7.4% of our DD cohort harbored a noncoding BCR breakpoint that disrupted one of these six TADs. We also defined a subset of core genomic features that, when considered together, can aid in the interpretation of DD-associated LRPEs throughout the genome.

## RESULTS

### International aggregation, sequencing, and genome-wide analyses of BCRs in 406 DD cases and 304 controls

We have previously shown that chromosomal rearrangements that appear balanced at cytogenetic resolution can involve extensive complexity ranging from multiple cryptic breakpoints to balanced chromosomal shattering, or chromothripsis, at sequence resolution.^11,15,34^ Here, we focused analyses on the most interpretable classes of BCRs by aggregating a cohort of 710 unrelated individuals harboring a “simple” BCR (*i*.*e*., breakpoints at two genomic positions without significant imbalance or additional complexity) initially identified by cytogenetic methods and subsequently resolved using either short-insert or long-insert WGS (**Fig. 1A**; **Supplementary Fig. 1**; **Supplementary Methods**). This cohort included 406 cases diagnosed with a DD or congenital anomaly in which a BCR was confirmed to have arisen *de novo* or segregate with phenotype and 304 unaffected control adults with no early-onset pediatric phenotype (see **Fig. 1B** and **Supplementary Table 1** for complete descriptions). Over 55% of the BCR breakpoints have not been previously published. As a comparison for these 710 empirically-identified BCRs (*n =* 1,420 breakpoints; **Supplementary Table 2**), we also generated a set of 30,400 simulated BCRs under the null hypothesis that breakpoints should be randomly distributed throughout the genome. These simulated BCRs were randomly sampled *in silico* from the genome while matching properties of the 304 BCRs empirically identified in controls, including structural variant (SV) type, inversion size, as well as excluding N-masked regions known to be inaccessible to short read alignments (**Supplementary Methods**).^35,36^

**Fig. 1.**
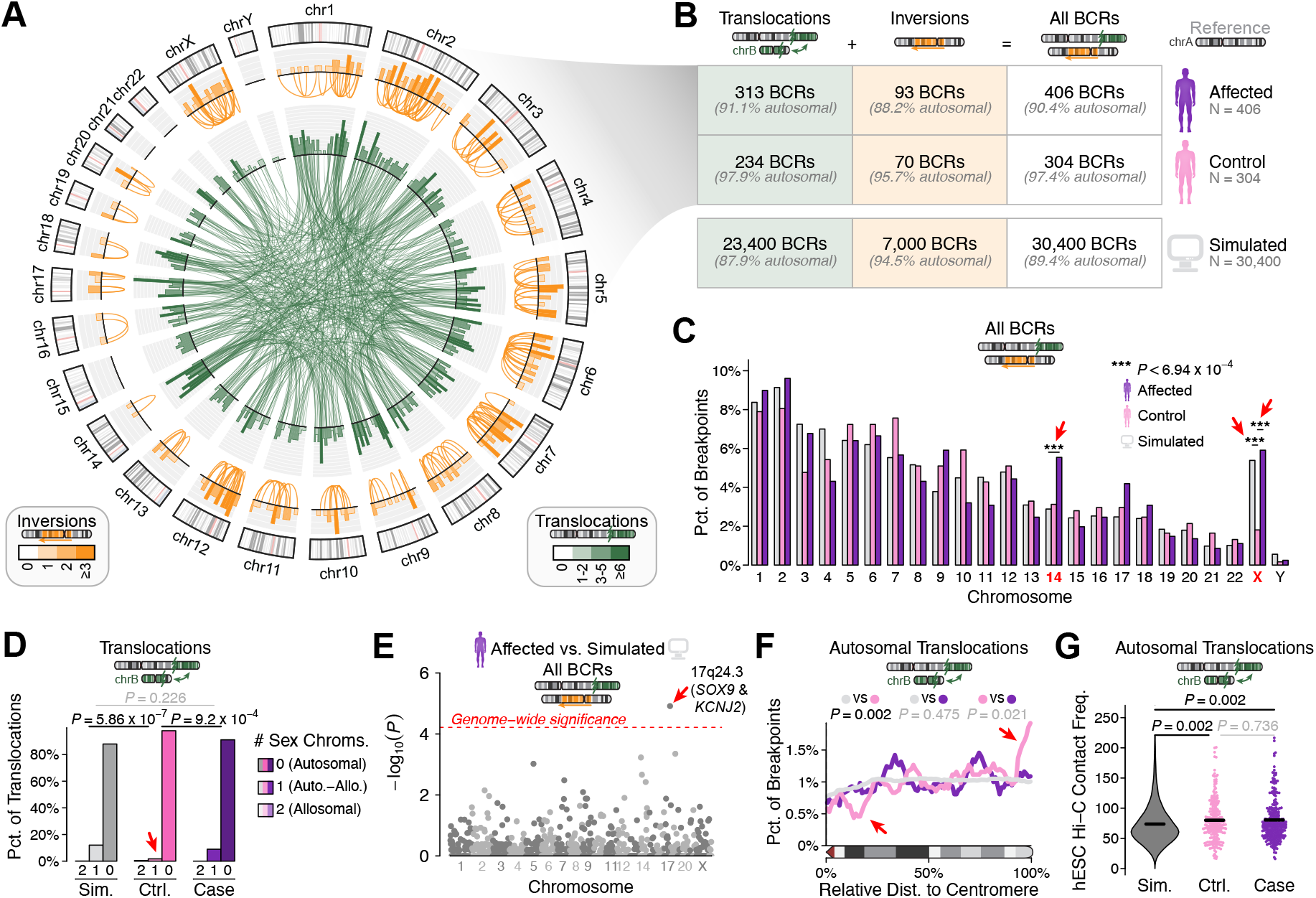
The properties of BCRs in the healthy human germline and DD cases. (**A**) We mapped the breakpoints of 710 simple (*i*.*e*., two-breakpoint) BCRs that were originally detected with cytogenetic methods. Here, we provide the genome-wide BCR breakpoint density in 10Mb windows per chromosome. (**B**) These 710 BCRs were identified in the genomes of 406 individuals affected by DDs and 304 unaffected controls. For purposes of comparison, we also generated 30,400 synthetic BCRs *in silico* by resampling the distribution of control BCRs 100 times randomly from the genome. (**C**) BCR breakpoints were distributed across the chromosomes as expected between affected, control, and simulated subsets except for chromosome 14, which exhibited a significant enrichment of breakpoints in affected samples, and chromosome X, which was depleted of breakpoints in controls. All comparisons were Bonferroni-adjusted for 72 total tests. (**D**) Balanced translocations involving at least one sex chromosome were significantly depleted in control samples compared to either affected samples or simulated null expectations. (**E**) We conducted association tests per cytoband for BCR breakpoints in affected samples vs. simulated null expectations. After correcting for 862 independent tests, just one cytoband was significant at the Bonferroni-adjusted threshold of *P* ≤ 5.8×10^−5^): 17q24.3, which corresponds to a locus with well-described LRPEs in DDs involving *SOX9* and *KCNJ2*.^24,25^ (**F**) The distribution of autosomal BCR breakpoints in controls across a “meta-chromosome” arm (*i*.*e*., chromosome size-normalized position) was significantly different from those of affected samples or simulated null expectations, with controls exhibiting depletions near centromeres and enrichments very close to telomeres. (**G**) Loci corresponding to empirically identified autosome-autosome translocation breakpoints (*i*.*e*., those sequenced in affected or unaffected genomes) contacted each other in 3D within the nucleus of human embryonic stem cells 1.08-fold more frequently than expected vs. simulated null expectations.

We first sought to understand the global patterns of BCRs throughout the genome by comparing the rates of breakpoints per chromosome between cases, controls, and simulations (**Supplementary Fig. 2**). We found that BCR frequency was approximately proportional to chromosome length with two exceptions: translocations were enriched on chromosome 14 in DD cases (*P =* 5.8×10^−6^ for cases vs. random simulations) and were depleted on chromosome X in controls (*P* = 1.4×10^−6^ for controls vs. random simulations) (**Fig. 1C** and **Supplementary Fig. 2**). The distribution of breakpoints across chromosome 14 did not appear to cluster in any particular location (**Supplementary Fig. 3**) and we did not find any features (*i*.*e*., compartment state, replication timing, recombination frequency, or gene disruption) that could account for the enrichment. In contrast, when we subset chromosome X, we observed that both cases and controls were 2.7-fold to 5.3-fold depleted for breakpoints on the q-arm (**Supplementary Fig. 2 and 3**). The Xq depletion in controls may be partly explained by the exclusion of males with oligo/ azoospermia and females with premature ovarian failure given their known association with X-autosome translocations.^37,38^ However, most of our DD cases were too young to be assessed for infertility, thus the Xq depletion in cases is unlikely to be related to a similar ascertainment bias. Overall, we identified that most (65%; 20/31) translocation breakpoints on chromosome X localized to the p-arm (length=58.6 Mb), which exhibited a 6.9-fold enrichment of case vs. control breakpoints (*P* = 0.002). Interestingly, 90.0% (18/20) of the Xp translocations identified in DD cases were found in females. Finally, controls were depleted for translocations involving either sex chromosome: just 2.1% of control translocations involved either chromosome X or Y, which was significantly less than translocations in DD cases (8.9%; *P* = 9.2×10^−4^) or randomly simulated BCR carriers (12.1%; *P* = 5.86×10^−7^) (**Fig. 1D**).

Our assessments of BCR breakpoint distributions per chromosome led to two additional discoveries. First, a single cytoband on chromosome 17 was significantly enriched for BCRs in cases (*P* = 1.2×10^−5^ vs. random simulations) and surpassed a genome-wide significance threshold adjusted for all 862 cytobands tested across all chromosomes (**Fig. 1E**; **Supplementary Fig. 4**).^39^ This cytoband, 17q24.3, matches the location of a well-described pathogenic LRPE in DDs and congenital anomalies caused by SVs altering the local TAD organization and dysregulating *SOX9* and *KCNJ2*.^24,25^ Second, after transforming the position of each breakpoint into a percentile relative to the length of its corresponding chromosome arm (*i*.*e*., meta-chromosome), we found that translocation breakpoints in controls were biased towards the most distal ends of chromosomes (Kolmogorov-Smirnov test; *P* = 0.002 for control vs. simulation and *P* = 0.021 for control vs. cases; **Fig. 1F**; **Supplementary Fig. 5**). For example, control BCR breakpoints were roughly three-fold enriched within the terminal 2% of each chromosome arm: 2.7-fold vs. DD cases and 3.3-fold vs. random simulations. This might suggest that translocations occurring near telomeres–which do not rearrange most of the affected chromosome–are more likely to be tolerated in the general population without leading to severe disease.

We also sought to identify genomic features that predispose to BCR formation by annotating all BCR breakpoints with features relating to chromosome maintenance (*e*.*g*., recombination rate, replication timing), chromatin accessibility, sequence context (*e*.*g*., repetitive elements, sequence homology), and three-dimensional (3D) nuclear organization (*e*.*g*., Hi-C contact frequency, nuclear compartment state).^40^ Most features showed no significant differences from expectations after correcting for multiple testing. One feature of note was that our empirically-observed translocations (*i*.*e*., those sequenced in DD cases and controls) were slightly more likely to form between pairs of chromosomes in close proximity to each other in 3D within the nucleus than predicted from simulated breakpoints that did not account for this biological organization (1.08-fold increase; *P* = 0.002; **Fig. 1G**). This result was true in a fetal lung fibroblast cell line (IMR90) and replicated in a second dataset derived from embryonic stem cells.^40^ These findings might suggest a weak influence on the formation of BCRs between chromosomal regions that co-localize within the nucleus, as suggested by analyses of tumor genomes and cytogenetic data from germline BCR carriers.^41^

### BCRs in DD cases are strongly enriched for direct disruption of established disease genes

Previous studies have demonstrated that BCRs confer substantial risk for DDs through direct disruption of haploinsufficient, developmentally critical genes.^8,11,15^ However, the absence of matched cohorts of unaffected control BCR carriers has historically hindered the quantification of disease risk contributed by gene-disruptive BCRs. Here, we annotated all BCR breakpoints for direct gene disruptions using Gencode v19 and compared the frequency of gene-disrupting autosomal BCRs between DD cases, controls, and random simulations.^42^ Most BCRs disrupted at least one protein-coding gene and there was no difference between cases and controls (68.1% of DD cases and 67.6% of controls) or expectations from random simulations (69.0% expected; **Fig. 2**). We further subdivided protein-coding genes into four tiers based on the evidence for their association with disease (**Supplementary Table 3**). Briefly, these included genes associated with dominant DDs (*i*.*e*., Tier 1, *n* = 812), genes associated with all other diseases (Tier 2, *n* = 3,129), mutationally constrained genes with no prior disease association (Tier 3; *n* = 1,257), and all remaining protein-coding genes (Tier 4; *n =* 15,188). We found a strong enrichment of cases with BCR breakpoints directly disrupting Tier 1 genes compared to controls (21.3% of cases versus 3.4% of controls; odds ratio [OR] = 7.45; 95% confidence interval [CI] = 3.74-16.50; Fisher’s exact test; *P =* 1.60×10^−12^) and compared to simulations (21.3% of cases vs. 6.6% of simulations; OR = 3.67; 95% CI = 2.80-4.76; *P =* 2.06×10^−18^), but not for Tiers 2-4.

**Fig. 2.**
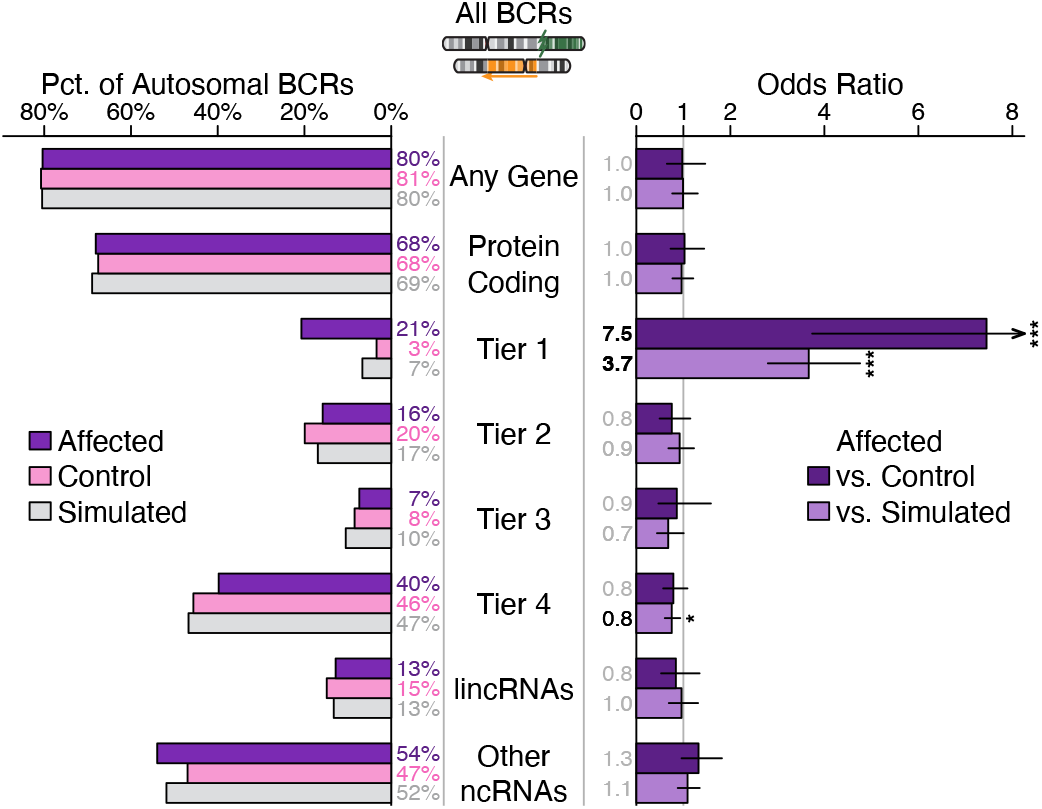
Risk conferred for DDs by BCRs disrupting genes and other transcribed loci. We annotated all BCR breakpoints for predicted overlap with protein-coding and noncoding genes present in Gencode v19.^14^ Here, we further subset these BCRs based on the properties of the gene(s) disrupted at either breakpoint, including: any gene present in Gencode; protein-coding genes; “Tier 1” genes including those known to be associated with dominant DDs; “Tier 2” genes including all remaining disease-associated genes; “Tier 3” genes including all genes in the top decile of loss-of-function constraint^47^ but with no existing disease association; “Tier 4” genes including all remaining genes not captured in the preceding tiers; lincRNAs; all noncoding RNAs other than lincRNAs present in Gencode. For each subset of genes, we computed the rate of BCRs disrupting at least one qualifying gene between cases, controls, and simulated BCRs, and further computed the odds ratios of cases vs. controls and cases vs. simulated null expectations. Only BCRs disrupting Tier 1 genes were significantly enriched in cases vs. controls and cases vs. simulated BCRs after correcting for multiple comparisons.

Motivated by the strong association between disruption of dominant DD genes and BCRs in cases, we systematically searched for genes that were recurrently disrupted by BCRs in cases beyond expectations by conducting association tests for each autosomal gene (**Supplementary Table 4**). These analyses identified four protein-coding genes disrupted in at least three independent DD cases and none in controls (**Table 1**). Among these, just one gene, *TCF4*,^11,15^ surpassed a strict exome-wide significance threshold (disrupted in 1.6% of DD cases vs. 0.01% of simulated BCRs; *P =* 4.3×10-10; OR = 149.5). Haploinsufficiency of *TCF4* is the dominant genetic cause of Pitt-Hopkins Syndrome and has been associated with autism spectrum disorder (ASD) and broadly defined neurodevelopmental disorders (NDDs).^43,44^ The three remaining protein-coding genes did not reach exome-wide significance but had suggestive (*P* ≤ 0.005) evidence of association with DDs in our analyses. These included two established DD genes (*AUTS2, MBD5*)^45,46^ and one candidate DD gene, *CDK6*, which was disrupted in three cases that presented with developmental delay (*n* = 3), speech delay (*n* = 2), microcephaly (*n* = 2), and cardiac defects (*n* = 1). *CDK6* is highly constrained against damaging point mutations,^47^ is ubiquitously expressed across tissues,^48,49^ and encodes a cyclin-dependent kinase with major roles in skin, blood, and breast cancers.^50^ *CDK6* has also been associated with a recessive form of primary microcephaly,^51^ but has not been previously associated with dominant germline disease.

**Table 1.**
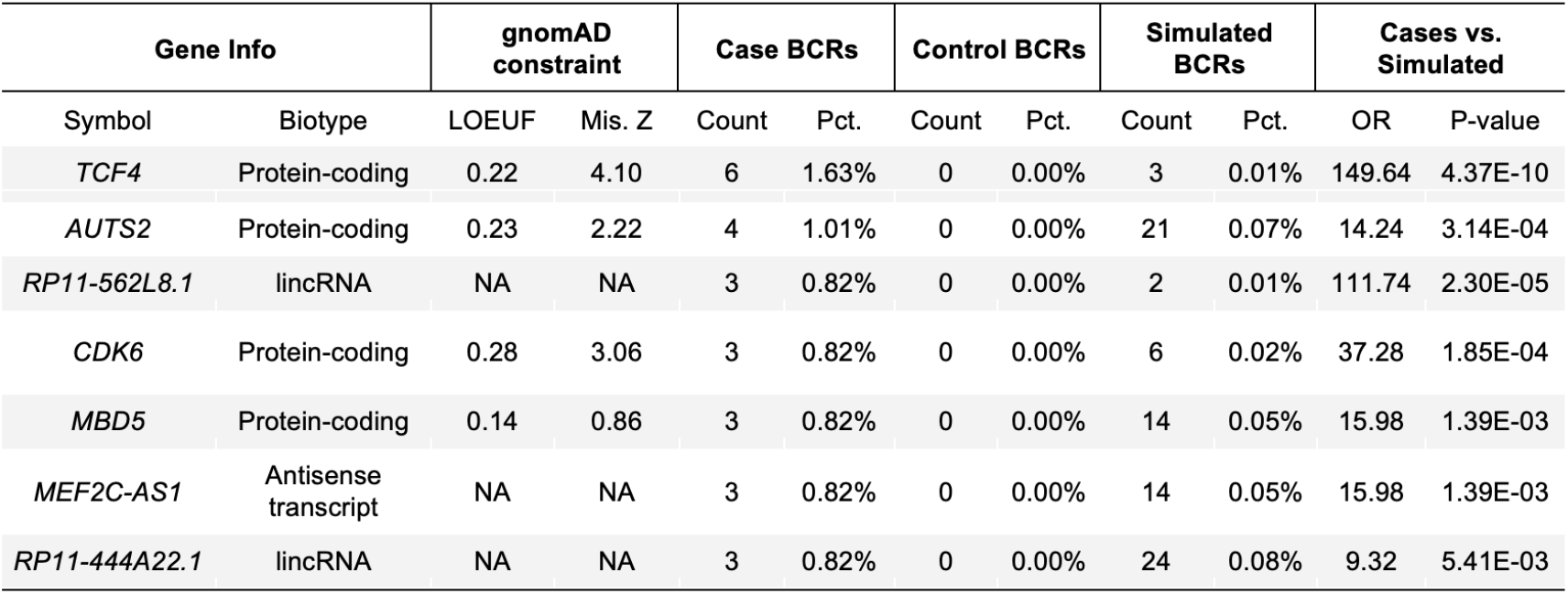
Genes recurrently disrupted by BCRs in DD cases. A list of seven genes that are disrupted by BCRs from ≥3 DD cases and zero controls. The type (“biotype”), constraint information,^47^ P value (for case versus control and simulated breakpoint comparisons, respectively), and odds ratio (OR) for each gene are also shown. Only one gene, *TCF4*, met a strict exome-wide significance threshold. LOEUF, loss-of-function observed/expected upper bound fraction; Mis., missense; Pct, percent; BCR, balanced chromosomal rearrangement.

### Pathogenic positional effects from disruption of three-dimensional chromatin structures

Our analyses identified an unambiguous, strong association between DDs and BCRs disrupting dominant DD genes; however, the majority of autosomal BCRs in DD cases did not disrupt a known disease gene (*n* = 289; 78.7%), and one-third (*n* = 113; 30.8%) did not disrupt any annotated protein-coding gene. We therefore considered three other models by which BCRs might confer DD risk through noncoding mechanisms based on: (i) their disruption of noncoding genes (*e*.*g*., lincRNAs), (ii) their linear distance from known disease genes, and (iii) the disruption of TADs containing known disease genes. We excluded all 78 (21.3%) cases and 10 (3.4%) controls with autosomal BCRs that directly disrupted a Tier 1 gene, which we reasoned would largely exclude the confounding influence of BCRs associated with pathogenic effects via direct gene disruption. We first tested whether direct disruption of noncoding genes could be responsible for pathogenic effects in DD cases and observed no difference in the fraction of cases versus controls that disrupted any subgroup of noncoding genes (**Fig. 2**). We next assessed whether pathogenic LRPEs could be predicted based on the absolute distance between disease genes and BCR breakpoints and observed no difference between DD cases and controls for proximity to any tier of genes (*e*.*g*., Tier 1 genes; Kolmogorov-Smirnov test; *P* = 0.159; **Fig. 3A-B**). These analyses confirm that linear distance to disease genes alone is insufficient to predict pathogenic LRPEs. However, when we tested the third model by comparing the fraction of cases to controls with a BCR breakpoint disrupting a TAD containing genes from each tier, we observed a significant effect for TADs containing Tier 1 genes (OR = 1.43; 95% CI = 1.68-3.18; Fisher’s exact test *P* = 0.033; **Fig. 3C-D**), supporting the role of 3D chromatin topology disruption in pathogenic LRPEs. Given these results, we next performed genome-wide analyses to define the TADs most strongly associated with DD phenotypes.

**Fig. 3.**
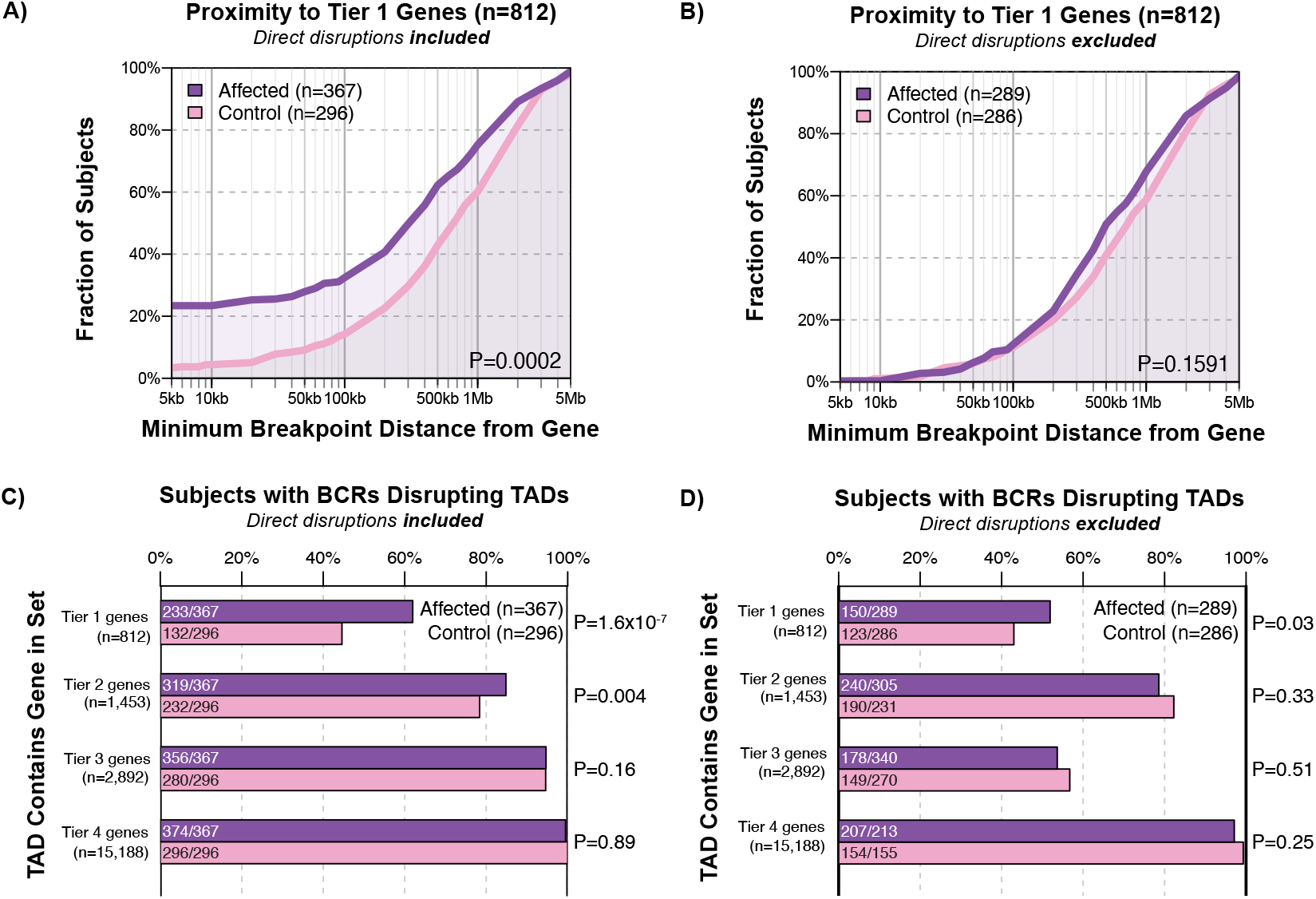
Disruption of TADs, and not proximity to known dominant DD genes, is predictive of LRPEs. (**A-B**) Fraction of cases and controls with an autosomal BCR breakpoint in proximity to a Tier 1 dominant DD gene (**Supplementary Table 3**) when direct disruption of Tier 1 genes are included (**A**) and excluded (**B**). P value corresponds to a two-sample Kolmogorov-Smirnov test. (**C-D**) Fraction of cases compared to controls with a BCR breakpoint directly disrupting TADs^40^ containing each of the four gene tiers when direct disruption of genes within that gene tier are included (**C**) and excluded (**D**).

We searched for specific TADs associated with risk for DDs by evaluating each autosomal TAD identified from a fetal lung fibroblast (IMR90) cell line^40^ for an enrichment of case BCRs against a Poisson null model fit to the distribution of control breakpoints. Overall, we identified 26 recurrently disrupted TADs with suggestive evidence for association with DDs based on a Benjamini-Hochberg false discovery rate (FDR) q ≤ 0.1 (**Fig. 4** and **Supplementary Table 5**). Five of these TADs surpassed a Bonferroni-adjusted genome-wide significance threshold of 2.2×10^−5^, including three known LRPE loci at *MEF2C, FOXG1*, and *SOX9* (**Fig. 5**).^11,14,52–54^ These five TADs also remained significant when we compared BCRs from DD cases against simulated breakpoints, suggesting that our models were not simply capturing rearrangement hotspots. Consistent with the finding that most TAD boundaries are tissue-invariant,^28,29,40^ tissue source had no impact on the genome-wide significant TADs (**Supplementary Fig. 6**). Given that we only removed cases and controls with a direct disruption of a Tier 1 gene, our TAD results represented a combination of true LRPEs, genic effects not previously associated with DDs, and other unknown mechanisms of disease etiology. For example, one of the genome-wide significant TADs was altered by three cases that all directly disrupted *CDK6*, a novel candidate DD gene identified from our exome-wide gene association analysis, suggesting that it likely represents a genic effect and not a LRPE.

**Fig. 4.**
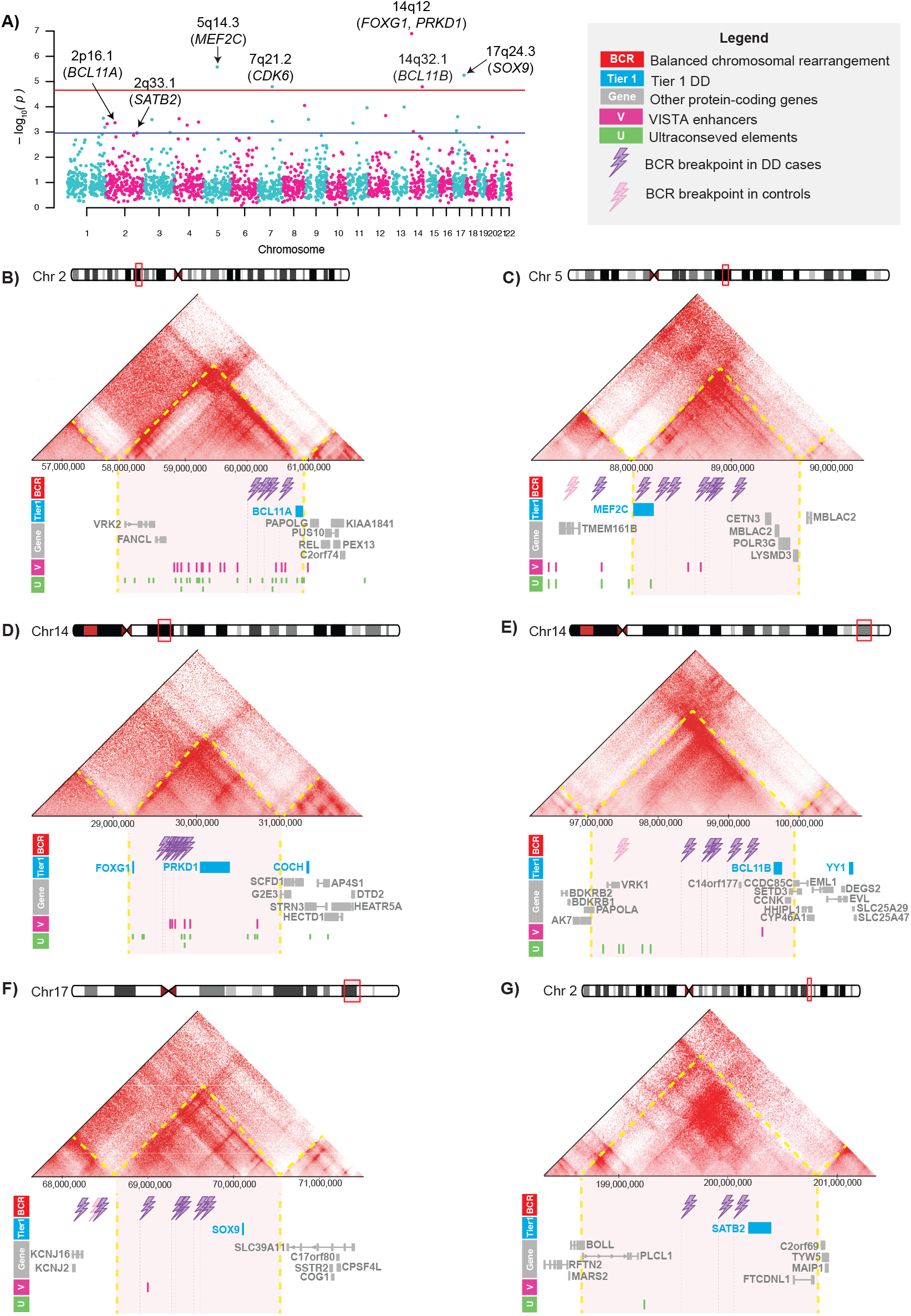
Genome-wide enrichment of TADs disrupted by BCR breakpoints from DD cases. (**A**) Genome-wide enrichment of BCR breakpoints in DD cases across 2,257 autosomal TADs.^40^ P values correspond to enrichments against a Poisson null model fit to the distribution of control breakpoints. The genome-wide significance threshold of 2.21×10^−5^ (denoted by the red line) was determined by correcting for the total number of autosomal TADs tested and the blue line represents the Benjamini-Hochberg FDR<0.1 cutoff. Known dominant DD genes (Tier 1) contained within each genome-wide significant TAD are reported in parentheses. The GM12878 Hi-C maps^30^ are shown for each of the six TADs significantly enriched for BCR breakpoints from DD cases: (**B**) 2p16.1 TAD containing *BCL11A*, (**C**) 5q14.3 TAD containing *MEF2C*, (**D**) 14q12 TAD containing *FOXG1* and *PRKD1*, (**E**) 14q32.2 TAD containing *BCL11B*, (**F**) 17q24.3 TAD containing *SOX9*, and (**G**) 2q33.1 TAD containing *SATB2*. The annotation tracks under the HiC maps include BCR breakpoints from DD cases and controls, Tier 1 DD genes (blue), all other protein-coding genes from Gencode v19 (gray),^42^ VISTA enhancers (pink),^62^ and UCEs (green).^63^

**Fig. 5.**
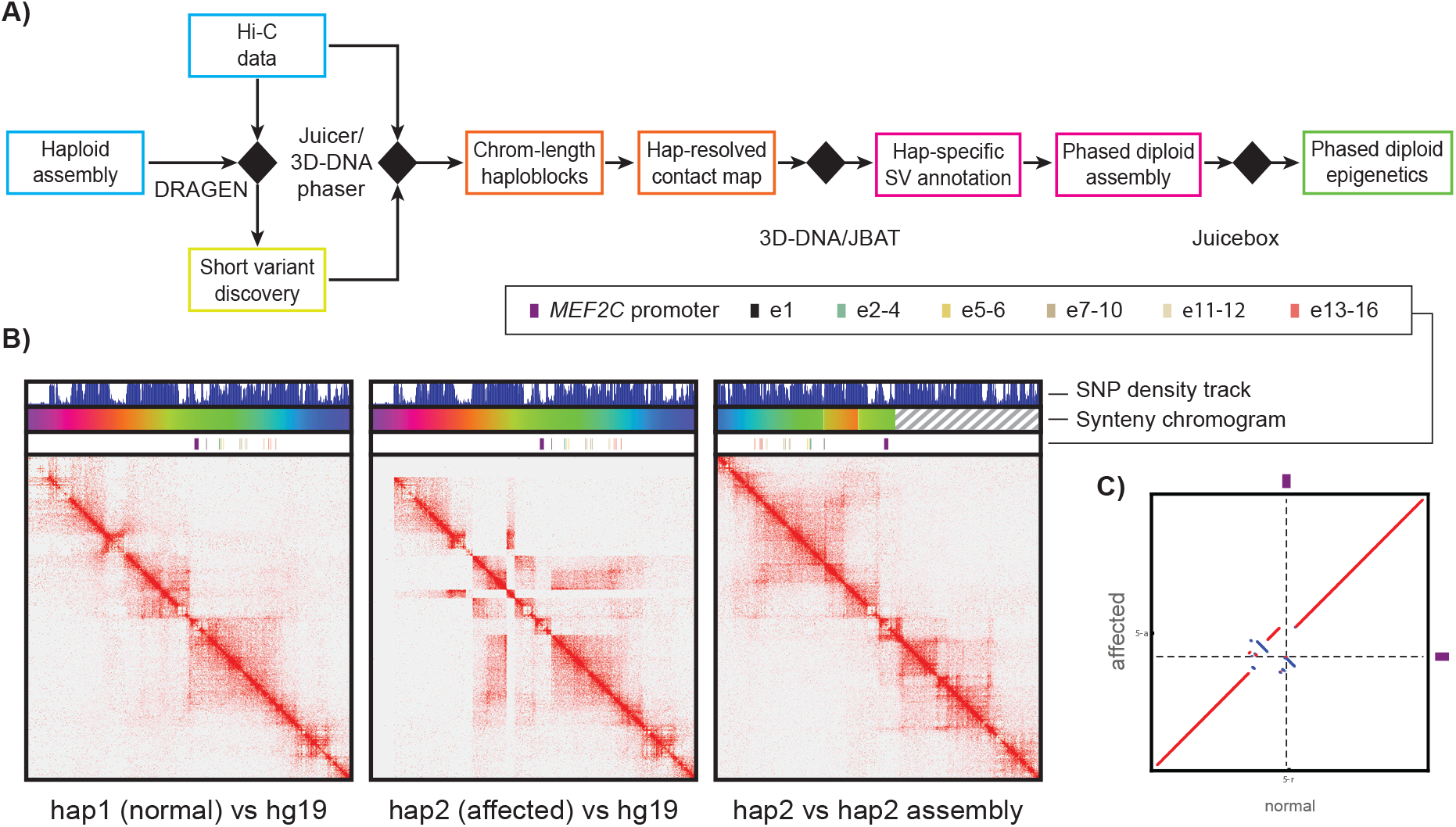
Hi-C analysis of DD case with complex BCR disrupting the TAD containing MEF2C. (**A**) The 3D resequencing pipeline starts by using Hi-C data to call short variants (SNPs and indels) against a haploid reference. In this paper we used the DRAGEN software,^79^ but similar results can be achieved with other publicly available variant callers such as GATK.^83^ We then use the Hi-C alignment data as generated by Juicer^84^ in conjunction with the 3D-DNA phaser to phase the variants and produce chromosome-length haploblocks. The phased variants enable the generation of molecule-specific contact maps, which in turn allow for molecule-specific annotation of SVs. Using the assembly tools from the 3D-DNA/Juicebox Assembly Tools ecosystem we then create assisted assemblies congruent with the annotated SVs and remap the contact data against the new reference to allow for phased diploid epigenetic analyses. (**B**) Molecule-specific Hi-C contact maps showing DNA-DNA interactions in the vicinity of the *MEF2C* promoter in LCLs derived from patient DGAP101: “normal” haplotype Hi-C data mapped to hg19 reference genome (left); haplotype with a chromothriptic chromosome 5 (middle, notice the numerous signal depletions along the diagonal corresponding to breaks and off-diagonal enrichments in the signal corresponding to fusion points); chromothriptic haplotype remapped against a reference that accounts for the chromotriptic rearrangements (right). The 1D tracks show the phased SNP density, highlight the syntenic regions between the three maps (rainbow colors are reserved for sequences in the vicinity of *MEF2C* in the “normal” reference, while hatching corresponds to sequences juxtaposed into the genomic segment of interest from elsewhere on chromosome 5 in the affected haplotype), as well as show the position of the promoter and 16 known enhancers^61^ in the ‘standard’ human reference as well as in the SV-corrected reference. (**C**) A dotplot of the whole chromosome 5 showing the correspondence between the affected and the normal molecules (100Kb synteny blocks are used, with direct synteny blocks colored red, and inverted blocks colored blue). The position of the *MEF2C* promoter is highlighted with dashed lines.

To identify candidate pathogenic LRPE loci among the 26 TADs with q ≤ 0.1, we required that each TAD: (i) be disrupted by a noncoding BCR breakpoint (*e*.*g*., does not disrupt a protein-coding gene) in at least 50% of cases contributing to the signal, (ii) contain a Tier 1 gene representing a plausible target gene, and (iii) have multiple cases disrupting the same TAD that present with phenotypes that are frequently observed in cases with direct disruption of the candidate target gene (**Supplementary Table 6**). This resulted in six candidate pathogenic TADs containing the known DD genes *SATB2, MEF2C, FOXG1, SOX9, BCL11B*, and *BCL11A* (**Fig. 4**). Supporting our statistical enrichments, five of the six significant TADs have been previously associated with pathogenic LRPE loci (*SATB2, MEF2C, FOXG1, BCL11B* and *SOX9*) based on individual case reports,^11,14,52,54-56^ suggesting that we are accessing *bona fide* LRPE signals. The novel candidate LRPE at 2p16.1 contains *BCL11A*, which encodes a zinc finger protein involved in the BAF SWI/SNF chromatin remodeling complex, and has been previously associated with an intellectual disability syndrome.^57^ All four of the cases with noncoding BCRs disrupting the TAD containing *BCL11A* presented with DD or ASD.^57^ Overall, these six TADs were disrupted by 30 cases (7.4%) and one control (0.32%; Fisher’s exact test; OR = 21.2; 95% CI 4.5-500.1; *P =* 7.12×10^−7^), suggesting that they represent highly-penetrant LRPE loci.

### Hi-C analyses of individuals with noncoding BCRs disrupting the TAD containing MEF2C

We previously implicated the 5q14.3 locus as a putative pathogenic LRPE with *MEF2C* as the target gene based on a statistically significant enrichment of noncoding BCR breakpoints that all disrupted the same TAD containing *MEF2C* and observed down-regulation of this gene in multiple cases harboring the noncoding BCRs.^11^ Based on these data, we hypothesized that the disruption of 3D topological organization could represent the underlying mechanism for this LRPE. To functionally validate this hypothesis, we generated high-throughput chromatin conformation capture (Hi-C) data from lymphoblastoid cell lines from five cases harboring BCRs disrupting the TAD containing *MEF2C* and developed a 3D resequencing workflow (see **Supplementary Methods** and **Fig. 5A**) to facilitate analysis of the resulting data. The goal of this workflow was to use Hi-C datasets to: (i) identify single nucleotide polymorphisms (SNPs), small insertions and deletions (indels), and SVs, (ii) phase these variants onto chromosome-length haploblocks, thereby reconstructing the end-to-end sequences of each molecule, and (ii) use the resulting diploid assembly to generate homolog-specific 3D contact maps.

We applied this workflow to two simple (one inversion and one translocation) and three additional complex BCRs from prior studies with noncoding breakpoints disrupting the TAD containing *MEF2C*.^11,14^ Each rearrangement was genotypically distinct with different resultant derivative chromosomes. While complex BCRs were excluded from other aspects of this study, we included three in our Hi-C analysis because their impact on 3D genome organization has not been previously examined in a homolog-specific manner.^59,60^ Comparing the results of rearrangement detection using Hi-C to those using liWGS, we found that the 3D resequencing pipeline detected 92.7% (*n =* 51/55) of the breakpoints found by the combination of both methods. Three of the four breakpoints missed by Hi-C were short segments (<10kb) of DNA that had been rearranged and inserted into a new position. The missed breakpoints were visible in the Hi-C map but had not been identified by the computational analysis. Conversely, liWGS detected 96.4% (*n* = 53/55) of the Hi-C identified breakpoints, failing to detect a breakpoint associated with a short interval, as well as a 78kb deletion. Crucially, Hi-C was able to reliably order and orient the rearranged sequences on each homolog, even when a breakpoint was missed (**Supplementary Table 7, Supplementary Figs. 7A-E**). By contrast, it is challenging to reliably order and orient the rearranged sequencing using liWGS data alone if breakpoints are missed. Taken together, these results demonstrate that Hi-C can be used to robustly generate both homolog-specific sequences and architectural maps.

In all five BCR cases we examined, 3D resequencing via Hi-C also revealed significantly altered 3D organization of the rearranged homolog (**Supplementary Figs. 7A-E**). Moreover, we observed reduced frequency of contact between the *MEF2C* promoter and 16 distal enhancers^60^ (**Supplementary Table 8**), consistent with the dysregulation of *MEF2C* expression observed in the same cases. In the majority of the cases, the reduction in contact frequency appeared to result from the BCR greatly increasing the distance between the promoter and its enhancers (**Supplementary Fig. 7**). However, in one case with chromothripsis (DGAP101), the BCR breakpoints only had a modest effect on the linear distance between the *MEF2C* promoter and its enhancers. Instead, the promoter and enhancers were separated into distinct architectural domains through the creation of a new boundary, which likely prevented physical contact between the promoter and enhancers (**Fig. 5B-C**). The observed 3D remodeling suggests a reduced frequency of contact between sequences that influence *MEF2C* expression, providing a plausible explanation for how a noncoding BCR breakpoint can result in a DD phenotype through disruption of 3D genome architecture.

### Genomic features predict TADs associated with pathogenic LRPEs

Motivated by our discovery of multiple genome-wide significant LRPE loci in DDs, coupled with our validation of functional changes in *MEF2C* and alterations to the 3D organization associated with the putative 5q14.3 LRPE, we exploited this unique BCR dataset to identify additional features contributing to pathogenic TAD disruptions that could be used for future LRPE predictions. As described above, we demonstrated that the genic content of TADs alone is insufficient to unequivocally predict the pathogenicity of an individual BCR, as 43.0% of BCRs in controls disrupted a TAD containing a Tier 1 gene. We also found that 9.4% of all TAD boundaries encompassing a Tier 1 gene were overlapped by at least one large polymorphic deletion in the genome aggregation database (gnomAD),^61^ which excludes adults with a history of early onset developmental conditions. Thus, additional genomic features beyond the presence of disease-associated genes are required to predict TADs preferentially disrupted by BCRs in DD cases.

To identify genomic features that characterize TADs intolerant to disruption, we annotated all autosomal TADs^40^ in the genome with 54 features that can be broadly grouped into five categories: genes, *cis*-regulatory elements, primary sequence conservation, repetitive elements, and ‘other’ (**Figs. 6A** and **Supplementary Methods**). We defined 45 “positive” training TADs (disrupted by ≥2 BCR cases and zero BCR controls) and 261 “negative” TADs (disrupted by ≥1 BCR control and no BCR cases) and performed a univariate logistic regression for each of the 54 features, which identified 26 features at a FDR<0.05 (**Supplementary Table 9**). Next, given that many genomic features are highly correlated (**Supplementary Fig. 8**), we trained an elastic net regression on the positive and negative training TADs that included all 26 features from the univariate analysis and identified six features that were individually associated with case status after controlling for the effects of all other features: VISTA enhancers,^62^ ultraconserved elements (UCEs),^63^ transposon-free elements,^64^ TAD size,^40^ the presence of at least one Tier 1 gene, and primary sequence conservation^65,66^ (**Fig. 6B**).

**Fig. 6.**
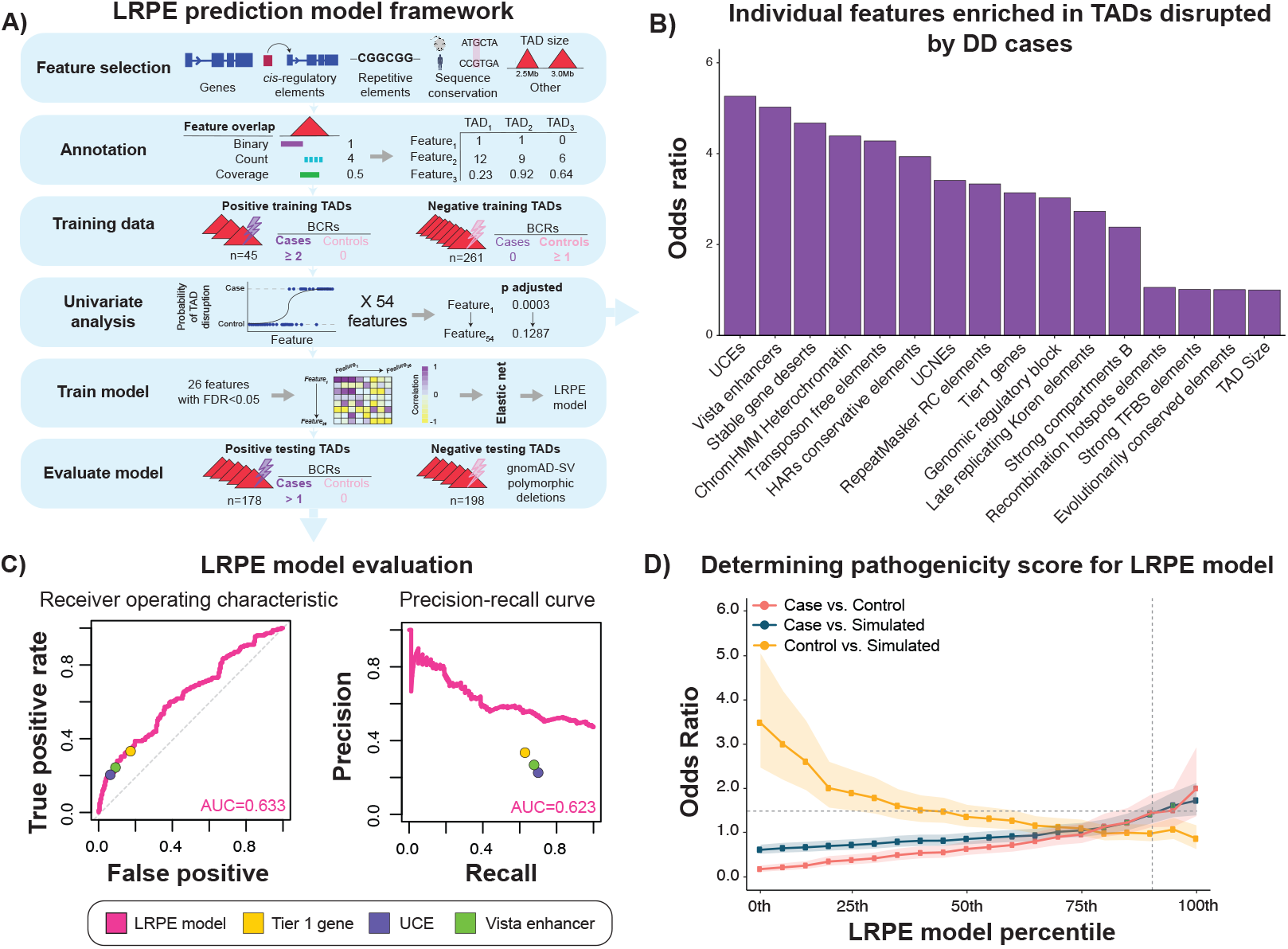
Identification of genomic features associated with pathogenic LRPEs. (**A**) An overview of the framework used to generate a model to predict pathogenic LRPEs across the genome based on disruption of 3D genome architecture. (**B**) Features identified to be potentially associated with TADs preferentially disrupted by BCRs from DD cases based on a BH-FDR<0.05 from a univariate logistic regression performed for each feature, ordered by effect size. (**C**) Evaluation of the LRPE model using an independent set of 372 TADs. Performance of TADs overlapping other common functional annotations are shown as a comparison. (**D**) To determine a cut-off score that could be used to identify TADs that are especially intolerant to disruption, we compared the fraction of cases to controls and simulated breakpoints that disrupt a TAD in each percentile and identified the inflection point at which a case enrichment (OR>1.5) begins to emerge.

We tested this model’s predictive accuracy on an independent set of 372 TADs (see **Supplementary Methods** for selection criteria) that were not used in our training data and determined that these six features alone were moderately predictive of LRPE pathogenicity (area under the receiver operating characteristics curve=0.633; **Fig. 6C**). We next defined a “LRPE pathogenicity” cutoff score of ≥0.43 (TADs ranked in the top 10th percentile from our model) based on the point in which case-control and case-simulation enrichments surpassed OR>1.5 (**Fig. 6D**). We demonstrated that 75.0% of previously established LRPE loci (**Supplementary Table 10**) in the human genome that were not represented in our training dataset surpassed this score, a highly significant enrichment when compared to all other TADs in the genome (Fisher’s exact test; *P =* 2.55×10^−5^). These orthogonal approaches collectively confirmed that this relatively simple six-feature regression model was able to prioritize TADs likely intolerant to disruption. While much larger cohorts will be required to power more sophisticated predictive models, these analyses demonstrate the potential of this approach to improve noncoding variant interpretation and shed greater light on the genomic features associated with noncoding mechanisms of disease.

## DISCUSSION

It has been well-established that *de novo* BCRs are associated with increased risk of congenital anomalies and a broad range of DDs,^2,4,6,10^ yet little is known about the pathogenic mechanisms through which this risk occurs outside of direct gene disruption. Using a large, aggregated cohort of cytogenetically-defined simple BCRs from which we derived sequence-resolved breakpoints, including several hundred BCRs from unaffected controls, our analyses reveal new insights into the mechanisms through which BCRs confer risk for DDs. These data identified an enrichment of BCRs impacting chromosomes 14 and Xp in DD cases, the latter of which was predominantly driven by X-autosome translocations in females. A previous cytogenetic study observed an association between telomeric breakpoints on the X chromosome, particularly at Xp22 and Xq28, and DDs in females with X-autosome translocations.^67^ Our analyses confirm and broaden these results by suggesting that this enrichment extends beyond Xp22 and likely encompasses the entire p arm. We further demonstrated that there is a seven-fold enrichment of BCR breakpoints that directly disrupt known DD genes in affected cases compared to controls, but that roughly 79% of DD BCR carriers cannot be explained by the direct disruption of currently recognized disease genes. We discovered TADs containing known DD genes that were recurrently disrupted by noncoding BCRs in DD cases far more frequently than expected by chance, suggesting that additional risk for DDs from BCRs are mediated through noncoding mechanisms, which represents an enticing area for future investigations.

These data implicate disruption of 3D chromatin domains as a mechanism likely mediating some of these noncoding effects. Our analyses identified six TADs enriched for BCR breakpoints in DD cases beyond what would be expected by chance. Notably, all three of the most significant TAD associations that exceeded genome-wide significant thresholds and matched previously established pathogenic LRPEs (*MEF2C, FOXG1*, and *SOX9*).^11,52,54^ The three additional LRPE loci are particularly compelling candidates given that they harbor well-known DD genes, *BCL11A, SATB2*, and *BCL11B*,^56,57,68–70^ and are disrupted by cases with phenotypes that match those seen in individuals with direct disruption of these DD genes. We note that, collectively, 7.4% of DD cases in this cohort harbor noncoding BCRs that disrupt these six LRPEs.

This result is also certainly an underestimate of the pathogenic impact of disruptions of 3D topology as it is restricted to only the six most prominent LRPE loci identified in our dataset. Many decades of disease gene discovery have biased our findings considerably towards identifying and prioritizing pathogenic gene disruptions, whereas much less is known about the molecular mechanisms, pathogenic processes, and genomic features of pathogenic noncoding variation. We anticipate that the fraction of BCR carriers associated with identifiable pathogenic LRPEs will continue to increase as future studies increase sample size and further clarify the essential features necessary for *cis*-regulatory disruption of disease-associated genes.

To investigate the potential underlying mechanism of LRPEs due to 3D topology disruption, we performed Hi-C analyses on five cases with noncoding BCRs that disrupted our top putative LRPE locus, the TAD containing *MEF2C*. While Hi-C has been performed on human cells from a small number of DD cases harboring SVs,^58,59^ none of these studies have explicitly isolated the impact of heterozygous SVs on 3D topology. This is largely due the fact that existing computational workflows do not effectively combine the phased *de novo* assembly of genomes with the discovery of SVs. Our 3D resequencing pipeline merged these tasks into a single workflow that was able to recapitulate the structure of each BCR detected by liWGS. In addition, the Hi-C analyses provided data on the 3D architecture at the 5q14.3 locus, revealing extensive disruption to the normal TAD structure of the region. The Hi-C analyses demonstrated that the 5q14.3 BCRs disrupted the physical contact between the *MEF2C* promoter and enhancers via two mechanisms, either by increasing the linear distance between them, or creating a new boundary that prevented the promoter from interacting with the enhancers. The creation of a novel boundary is consistent with a recent study that functionally dissected the TAD containing *SOX9*, one of our genome-wide significant TADs, and demonstrated that repositioning of the TAD boundary itself via inversion or insertion of novel CTCF sites created a new boundary that decoupled the *SOX9* promoter from its enhancers, resulting in downregulation of *SOX9* and abnormal phenotypic outcomes in mice.^24^ Additional Hi-C analyses of different SV classes at various loci will be critical for determining the full range of potential LRPE mechanisms, the relative prevalence of each, and what features predispose certain genomic regions to their pathogenic effects.

Our study further illustrates the complexity of interpretation of TAD disruption in human disease. We find that the disruption of TADs containing DD genes by control BCRs usually does not result in an appreciable disease phenotype, nor does the deletion of boudaries from these TADs as identified in population controls.^61^ This result refutes the utility of interpretation approaches that simply seek to match TAD disruption with the presence of a DD gene within the domain, particularly when assessing DD phenotypes that can be plausibly linked to hundreds of dominant disease genes throughout the genome. To systematically identify additional genomic features that were predictive of TAD intolerance, we leveraged our BCR cohort to build a model that prioritized six genomic features independently associated with TADs preferentially disrupted by DD cases. In addition to DD-associated (Tier 1) genes we also identified TAD size as being positively correlated with risk for DDs, which is consistent with developmentally regulated genes having complex *cis*-regulatory landscapes that likely occupy greater genomic space.^71^ We discovered an enrichment of primary sequence conservation and UCEs in TADs disrupted by BCRs from DD cases, which aligns with the report of an enrichment of UCEs in the vicinity of SVs associated with NDDs.^63^ Despite this initial progress towards identifying features associated with TADs intolerant to disruption, this model lacks sufficient predictive power to discriminate individual BCRs identified in cases from controls. However, we anticipate that larger sample sizes and improved noncoding functional predictions will eventually power increasingly sophisticated statistical models to aid in the clinical interpretation of noncoding BCRs. These data, together with our Hi-C analyses from the 5q14.3 locus, suggest that LRPEs are likely to be modified by the type of structural rearrangement as well as the complex interplay of genomic features within the TAD.

In conclusion, these data demonstrate that BCRs exert highly penetrant effects in DDs through both coding and noncoding mechanisms and that the disruption of 3D chromatin structures is associated with pathogenic LRPEs. We provide statistical evidence to support highly penetrant LRPEs at previously known and novel loci. Our feature selection analysis demonstrates that additional features within the TAD structure will also be critical for identifying novel LRPEs as well as provide insights into underlying molecular mechanisms through which this risk for disease occurs. The ongoing aggregation of population-scale datasets through international biobanks promises to further define the features associated with LRPEs and three-dimensional structures that are intolerant to disruption by SVs in the human genome.

## METHODS & SUPPLEMENTARY INFO

Detailed methods and supplementary information for this manuscript have been provided in a separate document, which will be linked directly from *medRxiv*.

## Data Availability

All reported breakpoints and case phenotypes have been made available in Supplementary Tables 1 and 2.

## ACKNOWLEDGMENTS

We thank the families and their clinicians for their participation in this study. This work was supported by grants from the The Danish National Research Foundation (WJC048 to N.T.), the Lundbeck Foundation (2007-1172; 2009-3999; 2010-6206; 2013-14290 to N.T.), the Danish Council for Independent Research (4183-00482B to N.T.), the National Institutes of Health (GM061354 to M.E.T, C.C.M, J.F.G., and E.L.; HD081256 to M.E.T.; MH115957 to M.E.T.; HD099547 to M.E.T.; HD091797 to M.E.T.; UM1HG009375 to E.L.A; RM1HG011016-01A1 to E.L.A.; HD090780 to S.L.P.S.; R00DE026824 to H.B.; GM007748 and DC012466 to B.B.C.; T32HG002295 to R.L.C.), the Simons Foundation for Autism Research (#573206 to M.E.T.), the National Science Foundation (NSF PHY-2019745 to E.L.A.; GRFP #2017240332 to R.L.C), Massachusetts General Hospital (Fund for Medical Discovery Fundamental Research Fellowship Award to C.L.), the Canadian Institutes of Health Research (Postdoctoral Fellowship to C.L.), the Welch Foundation (Q-1866 to E.L.A.), a McNair Medical Institute Scholar Award (to E.L.A.), a US-Israel Binational Science Foundation Award (2019276 to E.L.A.), the Behavioral Plasticity Research Institute (NSF DBI-2021795 to E.L.A.), the Investigator Grant Award Program (IGAP) BC Children’s Hospital Research Institute (to S.L.), the Czech Ministries of Health and Education (NU22-07-00165 and LM2018132 to Z.S.), the Estonian Research Council grant (PRG471 to K.Õ.), the Genome BC Grant (GR007838 to S.L.), the New Zealand eScience Infrastructure (to C.A.S.), the The IHC Foundation, Rutherford Discovery Fellowship administered by the Royal Society of New Zealand (to J.C.J.), the São Paulo Research Foundation (Fundação de Amparo à Pesquisa do Estado de São Paulo to A.C.S.F. and A.M.V.M.), the Council of Scientific & Industrial Research (to L.R.K.), the Fundamental Research Grant Scheme of the Malaysian Ministry of Higher Education (No. FRGS/1/2019/SKK08/UKM/02/9 to S.C.T.), the Polish National Centre of Science (No 2020/37/B/NZ5/00549 to M.K.), the funding support provided by Caroline Jones-Carrick and Collin Carrick (to H.G.K.), and the startup funding of the Qatar Biomedical Research Institute at Hamad Bin Khalifa University (to H.G.K.). We also thank Annemette Friis Mikkelsen and Bjarke Thomsen for their expert technical assistance; the Coriell Institute for Medical Research for providing biomaterials; and the Broad Institute Genomics Platform for generating a subset of the WGS data.

## AUTHOR CONTRIBUTIONS

**Study design:** C.L., M.M.M, R.L.C., M.C.B, O.D., H.B., C.C.M., E.L., J.F.G., P.J., E.L.A., I.B., M.E.T., and N.T. **Sample recruitment and data generation:** M.M.M, Z.D., M.B.R., L.N.P., A.S.F, Y.M., A.L.T., J.M.M.M., X.C., A.C., C.H., M.B., R.V.B., T.V., T.L., J.E., A.M.V.M., A.C.S.F., J.F.M., K.T.A., J.R.V., K.V.D.B., C.S., C.A., P.E., A.A.S., D.S., C.S.B., V.M.K., M.W., H.G.K., K.O., L.R., S.M., M.B., A.L.T., J.E., J.O., D.N., M.P., E.B., E.R.S., F.S., F.J.S., A.N., E.F., M.F., P.S., N.K.V., C.D.H., D.M.F., D.H.S., M.A.R.A., M.M.I., L.A., P.M.K., G.D.R., T.A.P.V., S.L., W.H, J.D., M.H., M.H., Z.S., I.V., T.D.H., R.S.M., C.F., L.B.O., B.S.G., M.L., J.P., C.P.R., S.J., N.E., E.S., C.M., K.G., A.D., U.R.D., R.S., F.L., O.Z., G.H., D.M., A.M., N.W.K., I.M.C., J.B.M., L.R.M., M.G., L.L., J.G., K.B.M.H., C.D.A.E., Y.A., J.R.B., X.L., J.L., A.S., C.H.H., L.N.K., J.T., K.L., R.H., R.G.S., C.A.S., J.C.J., B.L., A.T., B.N., E.M., O.A.C., E.S.W., T.K., R.E.P.A., S.G.T., A.K., A.R.H., S.B., S.L.P.S., N.Y., S.G., W.K.C., S.R., I.M., N.O., N.K.G., K.R.F., B.N.H., C.C.M., K.W.C., P.J., I.B., and N.T. **Individual case breakpoint identification:** M.M.M, M.C.B, C.L., M.B.R., H.B., Z.D., R.L.C., A.M.V.M., H.D.W., C.A., E.B., F.S., N.K.V., G.D.R., Z.T., L.B.O., C.A.S., J.C.J., K.N., S.G.T., S.O.S., B.T., K.W.C., P.J., I.B., S.C., S.C.T., A.Z., M.I.M., A.U., B.A. and N.T. **Aggregation data analysis and interpretation:** C.L., R.L.C., H.B., M.M.M, M.C.B, P.J., M.E.T., and N.T. **HiC data generation and/or analysis:** O.D., H.G., D.W., C.L., K.M.S., C.S.B., M.E.T., and E.L.A. **Generation of figures and writing of the manuscript:** C.L., R.L.C., M.M.M, M.C.B, O.D., H.B., H.G., K.M., E.L.A., M.E.T., and N.T. All authors revised the manuscript.

## COMPETING INTERESTS

M.E.T. receives research funding and/or reagents from Levo Therapeutics, Microsoft Inc, and Illumina Inc. E.L.A. receives in-kind support from IBM and Illumina Inc. M.M.M. and A.C.S.F. are employees of Illumina Inc. A.S.F. is employed by HERAX. All other authors declare no competing interests.

## Notes

### Author Declarations

All studies were approved by respective institutional review boards (IRB). Informed consent was obtained from all subjects or their legal representative for participation in the study when the IRB so required. The Mass General Brigham Human Research Committee (MGBHRC) Institutional Review Board (IRB) gave ethical approval for this work: Study Protocol 2013P000323, Genomic Studies of Human Neurodevelopment (September 07, 2018). Protocols undergo annual continuing review by MGBHRC IRB.

