## Supplementary Information for "Balanced chromosomal rearrangements offer insights into coding and noncoding genomic features associated with developmental disorders"

*Full list of authors is available in the main text*

### Table of Contents

**SUPPLEMENTARY FIGURES** 2

1. Genome-wide BCR breakpoint density in DD cases and controls 2
2. Enrichment of case BCR breakpoints per chromosome 3
3. Breakpoint positions per chromosome in cases and controls 4
4. Manhattan plots for all pairwise contrasts and SV types 5
5. Meta-chromosome density panels split by SV type 6
6. TAD enrichment across tissues 7
7. Phased Hi-C maps for cases with BCRs disrupting the TAD containing *MEF2C* 8
   7A. DGAP101 8
   7B. DGAP191 9
   7C. DGAP218 9
   7D. EB/0401 10
   7E. OL/2202 10
8. Correlation between genome features tested for association with LRPEs 11

**METHODS** 12

**SUPPLEMENTARY REFERENCES** 16

### SUPPLEMENTARY FIGURES

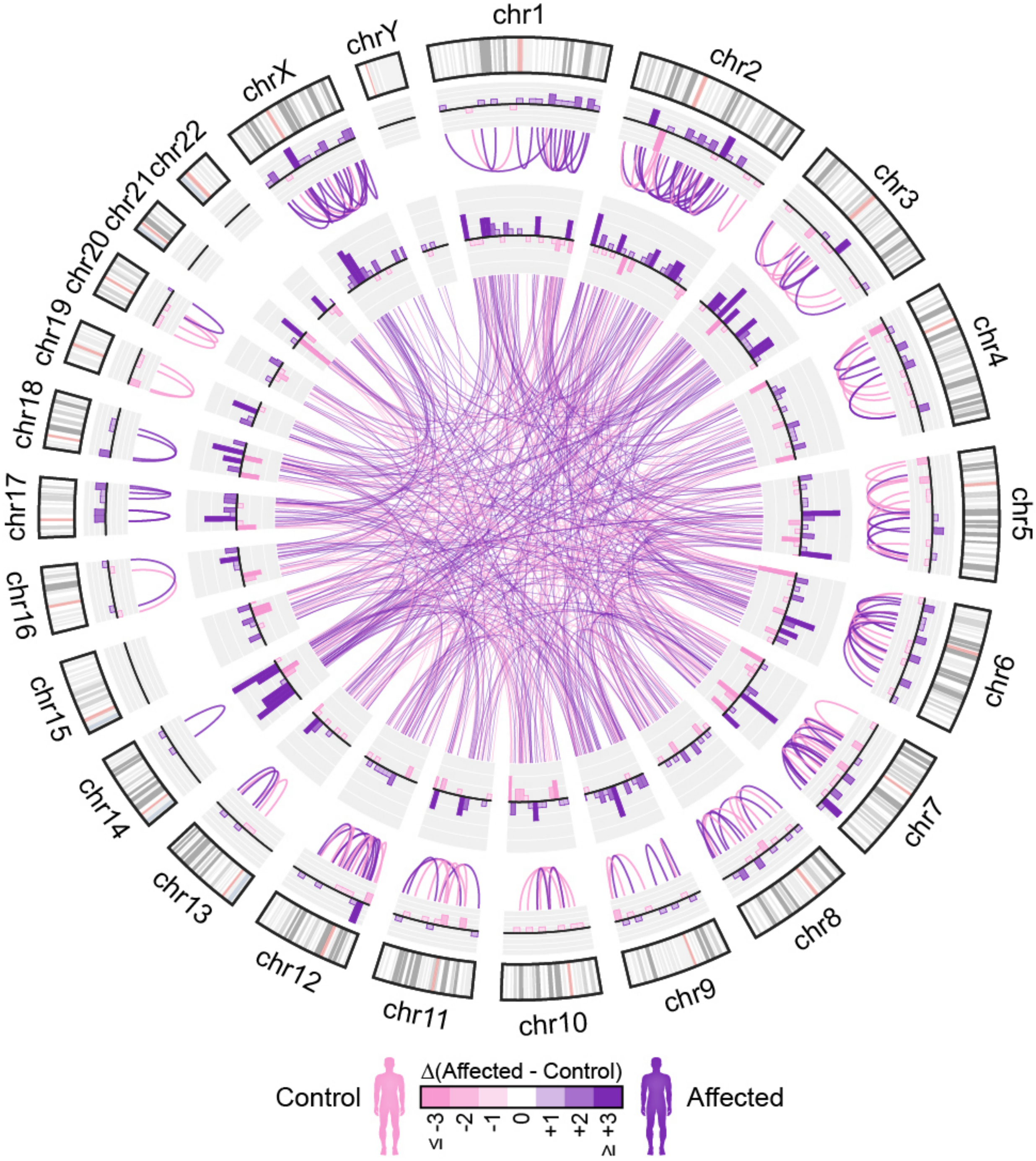


**Supplementary Figure 1. Genome-wide BCR breakpoint density in DD cases and controls.** Each BCR is denoted by a single link in purple (cases) or pink (controls). Histograms per chromosome correspond to the differential density of BCR breakpoints between cases and controls in 10Mb windows.

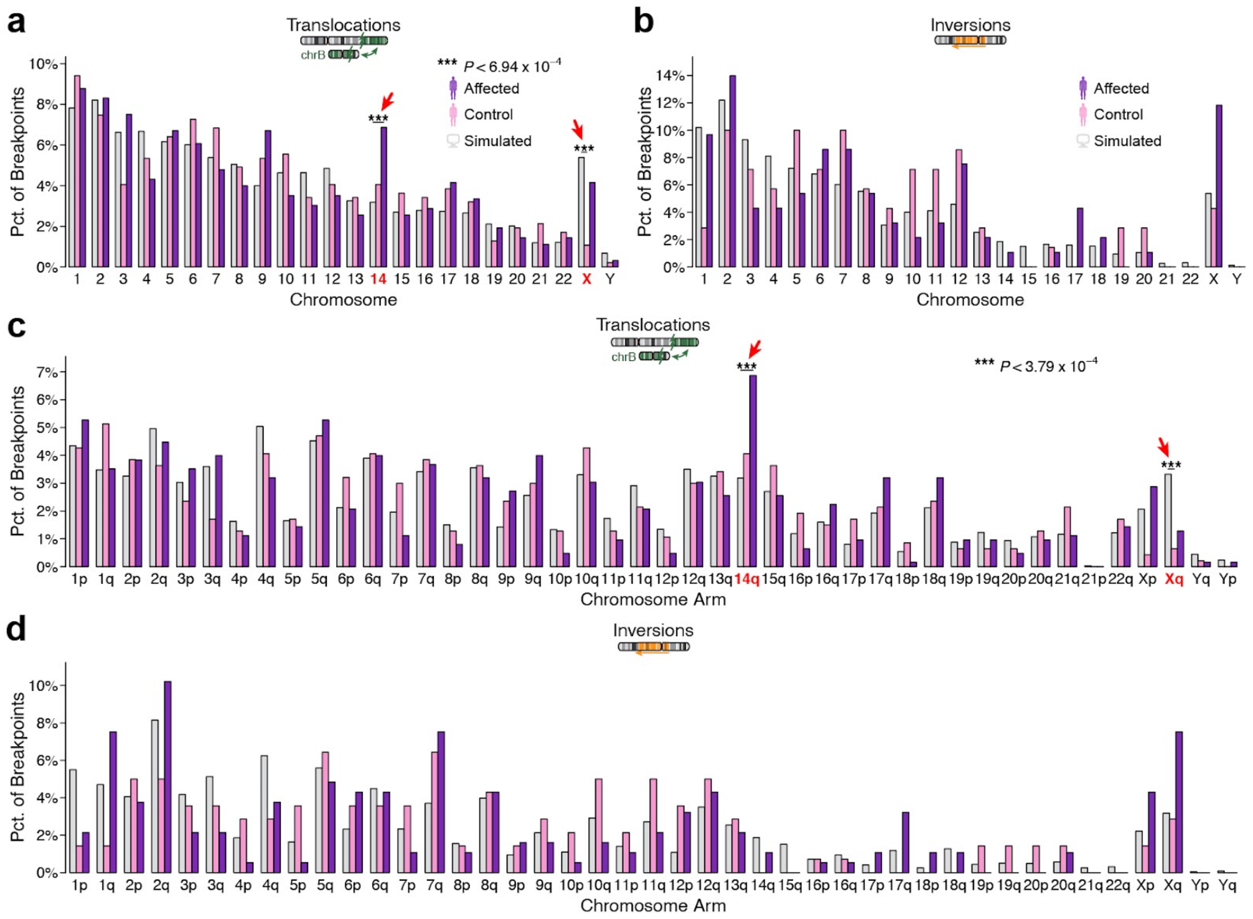


**Supplementary Figure 2. Enrichment of case BCR breakpoints per chromosome.** (a-b) Proportion of translocation (a) or inversion (b) breakpoints per chromosome for cases (purple), controls (pink), and simulated BCR carriers (grey). Triple asterisks indicate significant comparisons surpassing a Bonferroni-corrected threshold. (c-d) Proportion of translocation (c) or inversion (d) breakpoints per chromosome arm; formatting identical to panels (a-b).

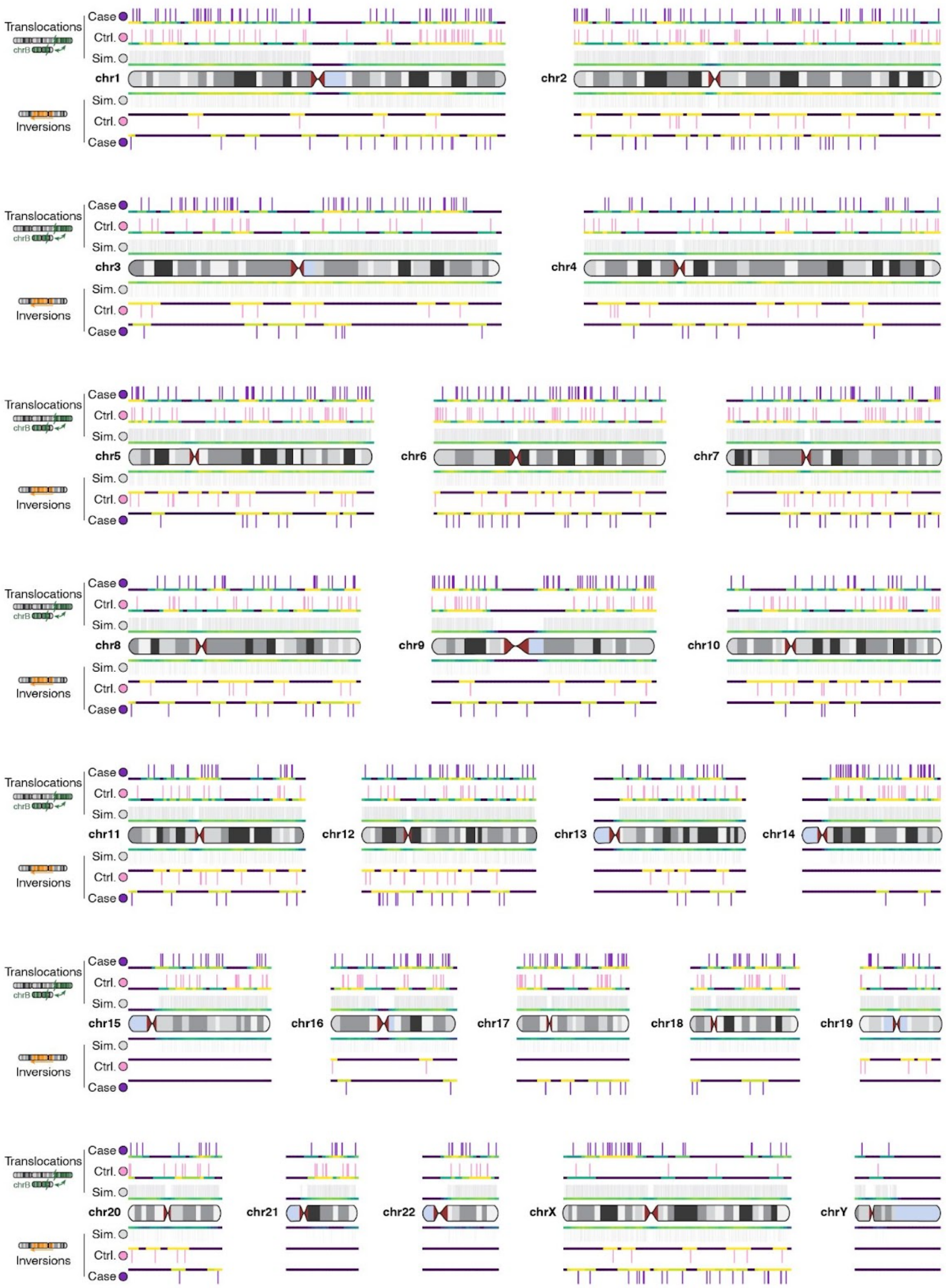


**Supplementary Figure 3. Breakpoint positions per chromosome in cases and controls.** Each vertical line represents the position of one breakpoint in cases (purple), controls (pink), or simulated BCR carriers (grey) for translocations (top panels) or inversions (bottom panels). Chromosomal regions shaded in light blue-grey are unalignable in the reference genome and were excluded when mapping breakpoints.

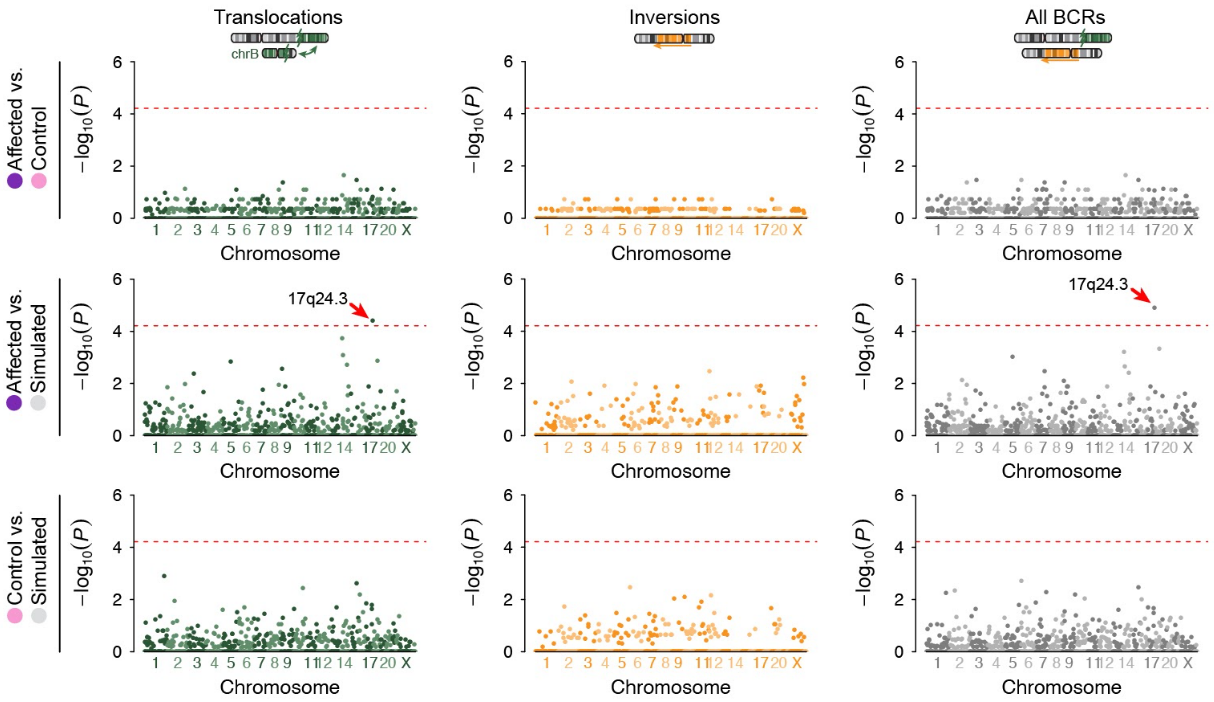


**Supplementary Figure 4. Manhattan plots for all pairwise contrasts and SV types.** Results from genome-wide BCR burden testing per cytoband. Each panel depicts a Manhattan plot of association results for a given phenotype contrast (rows) and BCA type (columns). Each point represents a single cytoband and horizontal dashed lines correspond to Bonferroni-corrected significance after adjusting for all cytobands tested.

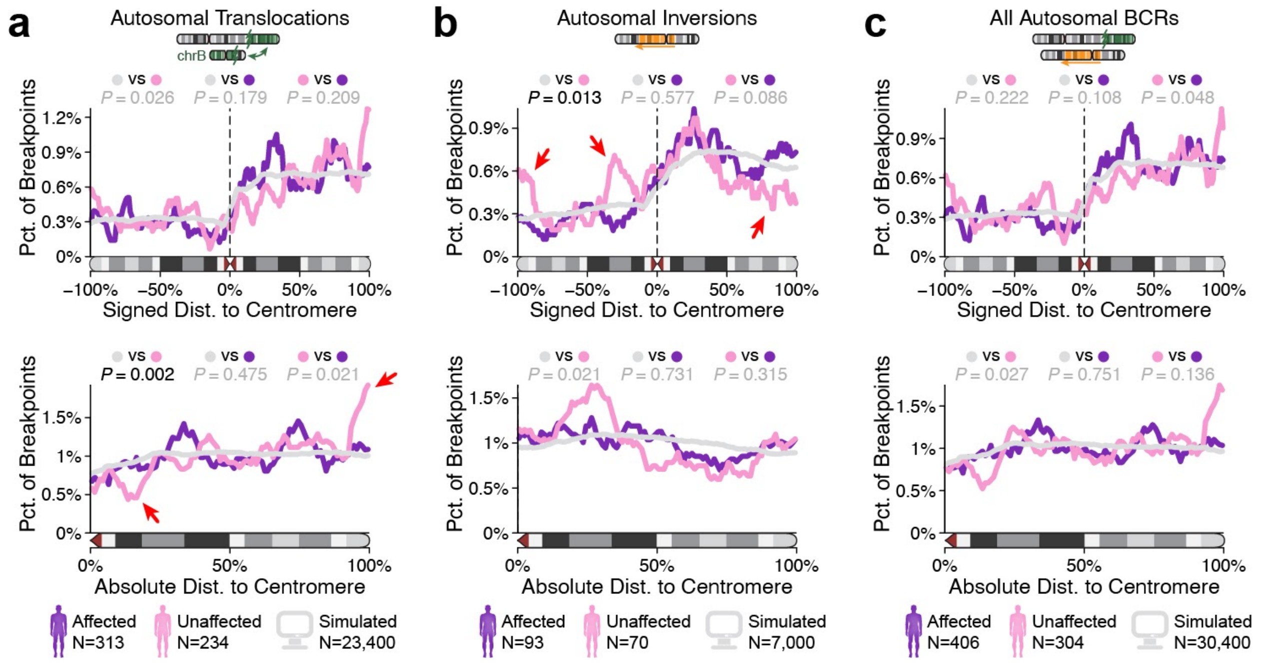


**Supplementary Figure 5. Meta-chromosome density panels split by SV type.** Autosomal BCR breakpoint densities normalized for chromosome length. Each panel provides the density of breakpoints from (a) translocations, (b) inversions, or (c) all BCRs in cases, controls, and simulated carriers after normalizing for chromosome arm length (see Methods). Data are presented as signed values in the top panel, where negative and positive values indicate P-arm and Q-arm breakpoints, respectively. Data are also presented as absolute values in bottom panels. Lines represent rolling smoothed averages of breakpoint densities per percentage increment on the X-axis, and densities were compared using Kolmogorov-Smirnov tests as described in Methods.

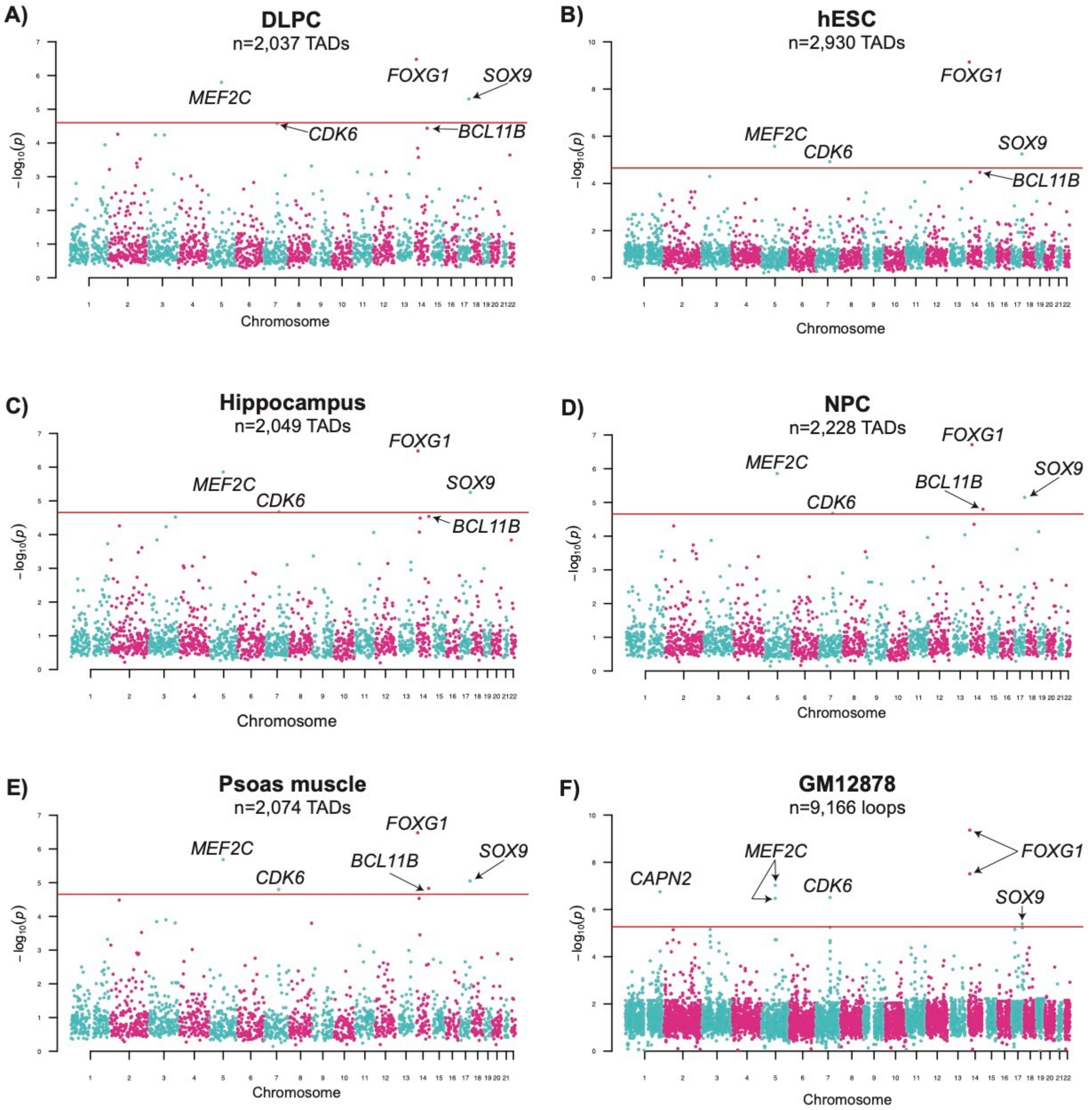


**Supplementary Figure 6. TAD enrichment across tissues.** Each panel represents TADs identified from a different tissue source that were tested for an enrichment of BCR breakpoints from cases, including the (a) dorsolateral prefrontal cortex,[1] (b) human embryonic stem cells,[2] (c) hippocampus,[1] (d), neural progenitor cells,[1] (e) psoas muscle,[1] and (f) loop boundaries from the GM12878 cell line.[3] The same approach that was used for the IMR90 cell line,[2] which is presented in the main manuscript, was applied to all tissues. The genome-wide significance threshold (denoted by the red line) was determined by correcting for the total number of autosomal TADs/loops for each tissue. The five genome-wide significant TADs from the IMR90 cell line are annotated based on their putative target gene.

**Supplementary Figures 7 A-E. Phased Hi-C maps for cases with BCRs disrupting the TAD containing *MEF2C*.** Molecule-specific Hi-C contact maps showing DNA-DNA interactions in the vicinity of the MEF2C promoter in LCLs derived from six cases with noncoding BCRs disrupting the TAD containing MEF2C. The "normal" haplotype Hi-C data mapped to hg19 reference genome (left); haplotype with carrying the rearrangement on chromosome 5; haplotype remapped against a reference that accounts for the rearrangements (right). The 1D tracks show the phased SNP density, the syntenic regions between the three maps (rainbow colors are reserved for sequences in the vicinity of MEF2C in the "normal" reference, while hatching corresponds to sequences juxtaposed into the genomic segment of interest from elsewhere on chromosome 5 in the affected haplotype), as well as show the position of the promoter and its 16 known enhancers[4] in the 'standard' human reference as well as in the SV-corrected reference. A dotplot of the whole chromosome 5 showing the correspondence between the affected and the normal molecules, where 100Kb synteny blocks are used, with direct synteny blocks colored red, and inverted blocks colored blue. The position of the MEF2C promoter is highlighted with dashed lines.

### Supplementary Figure 7A. DGAP101

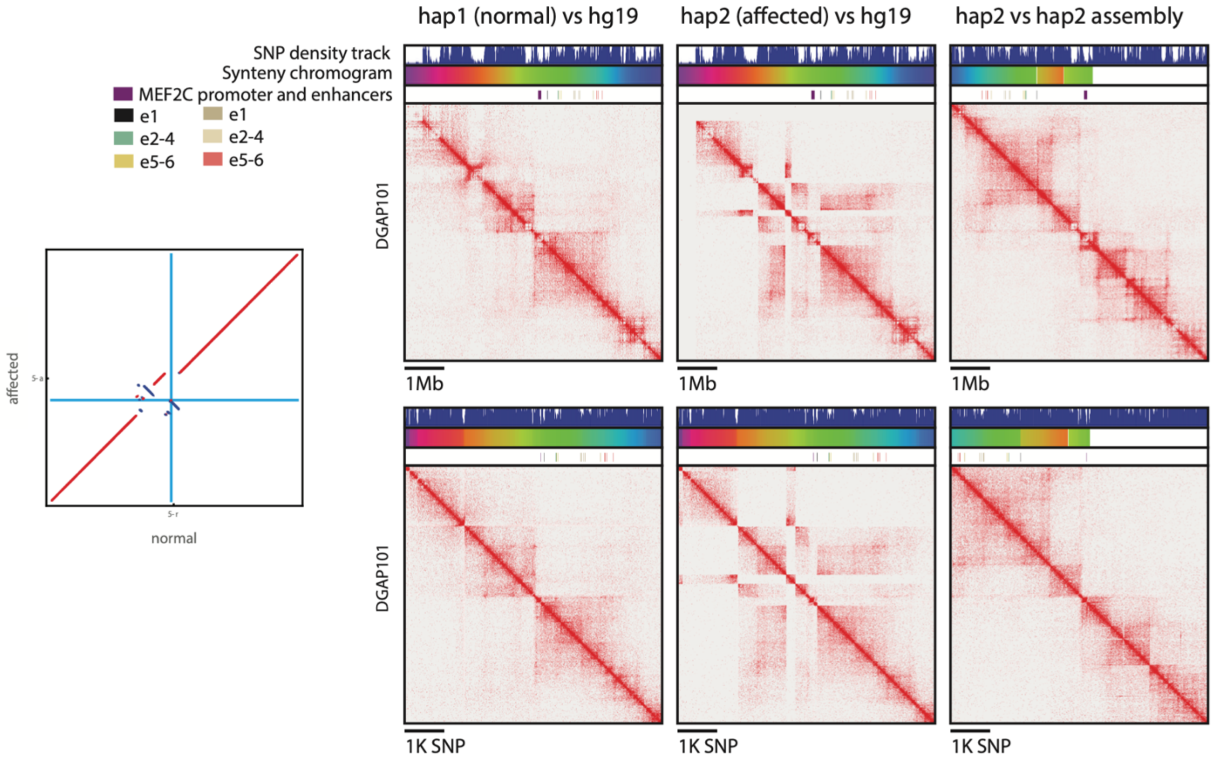

### Supplementary Figure 7B. DGAP191

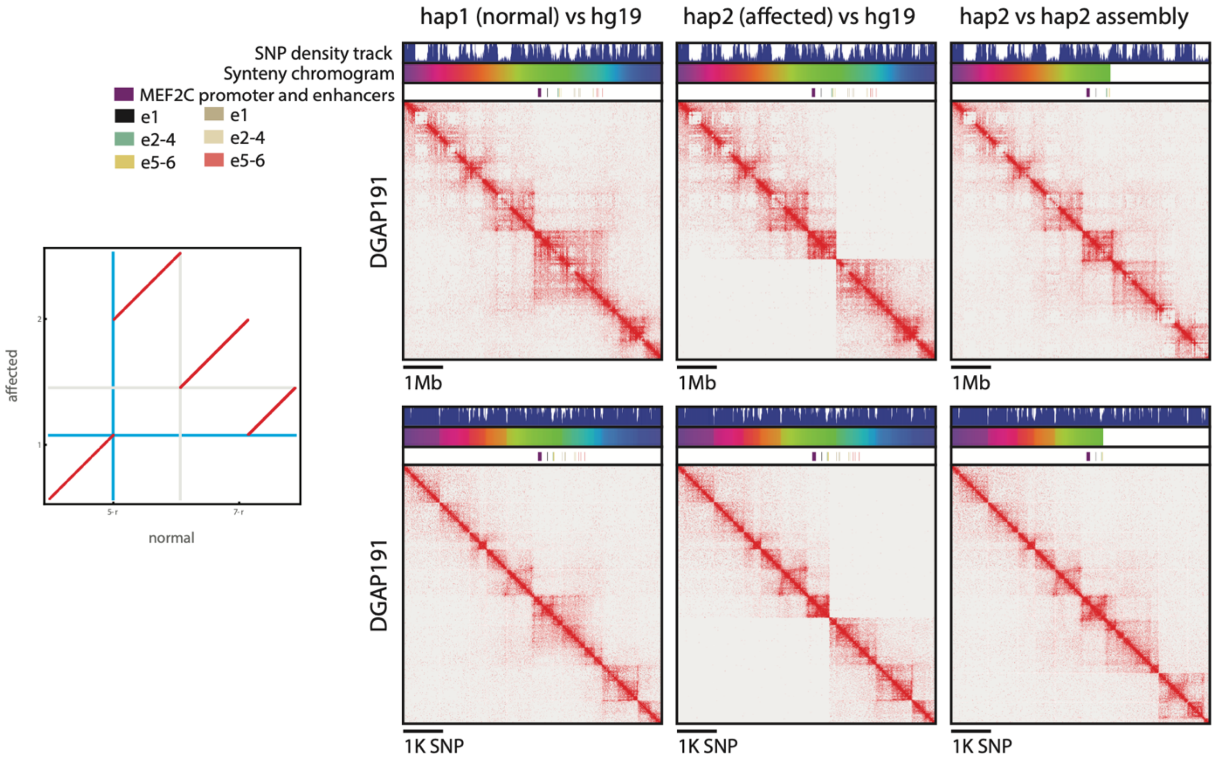


### Supplementary Figure 7C. DGAP218

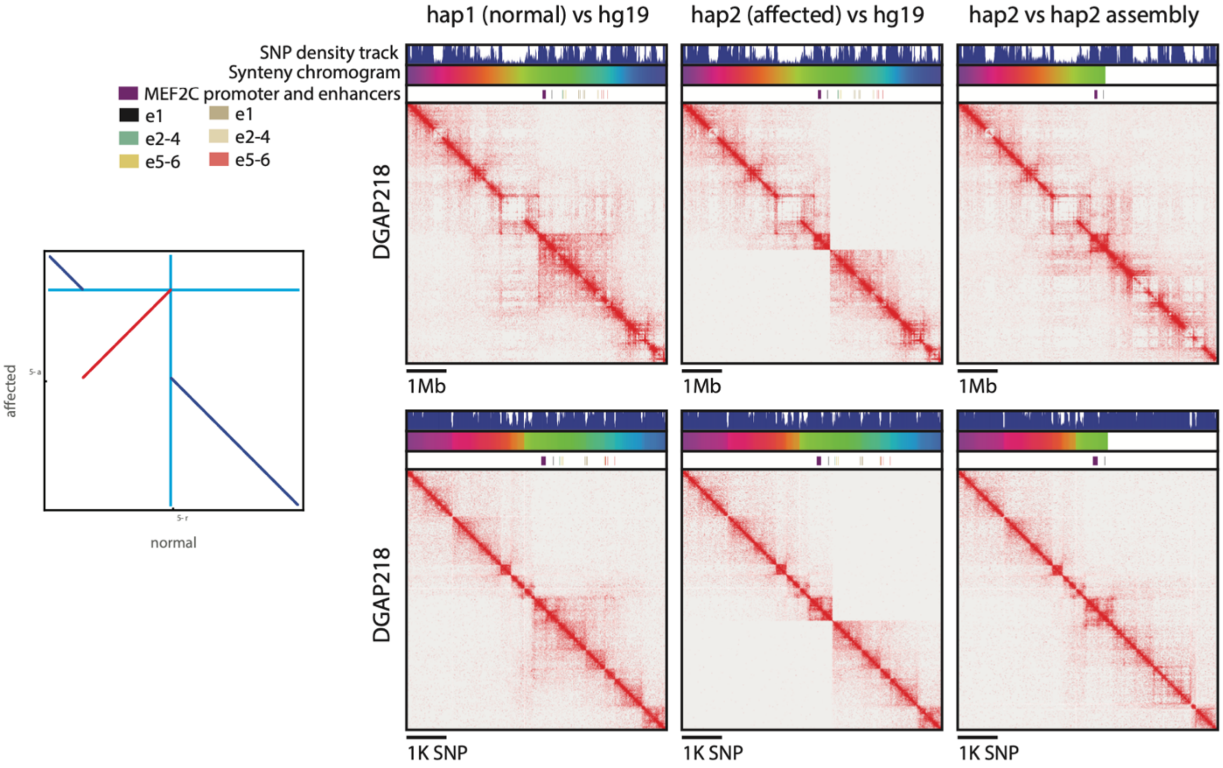

### Supplementary Figure 7D. EB/0401

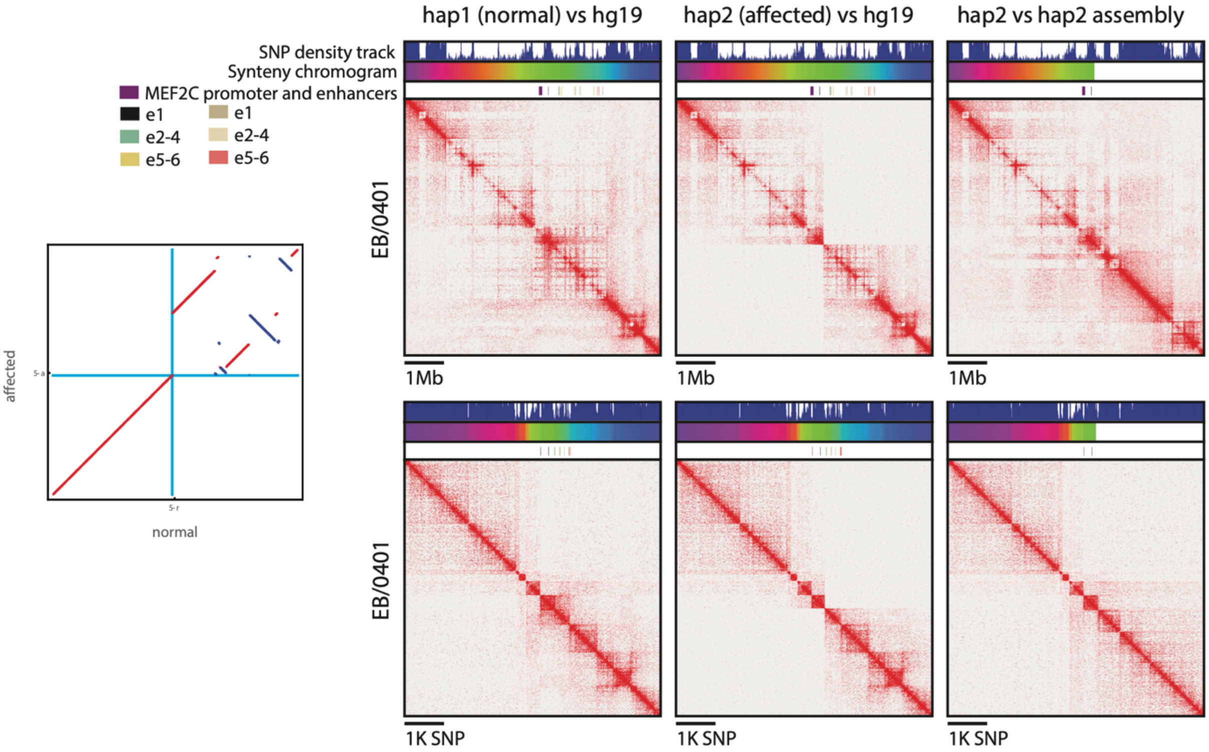


### Supplementary Figure 7E. OL/2202

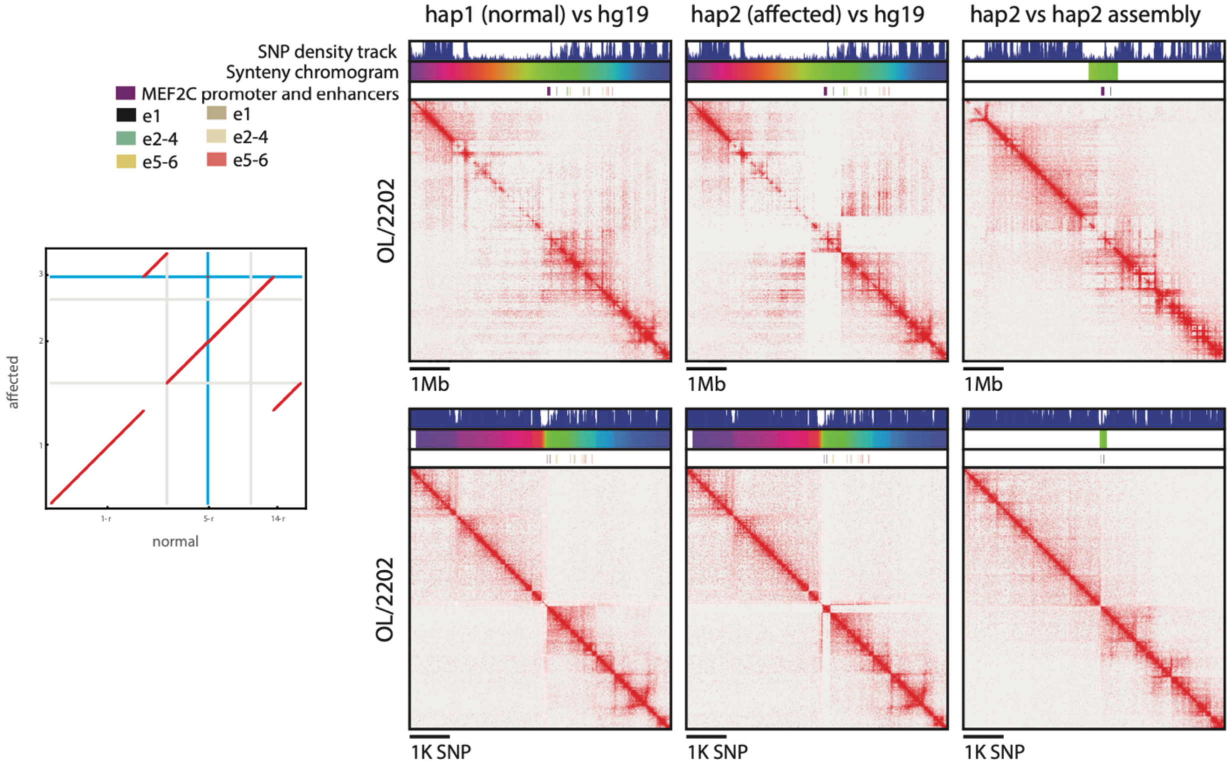

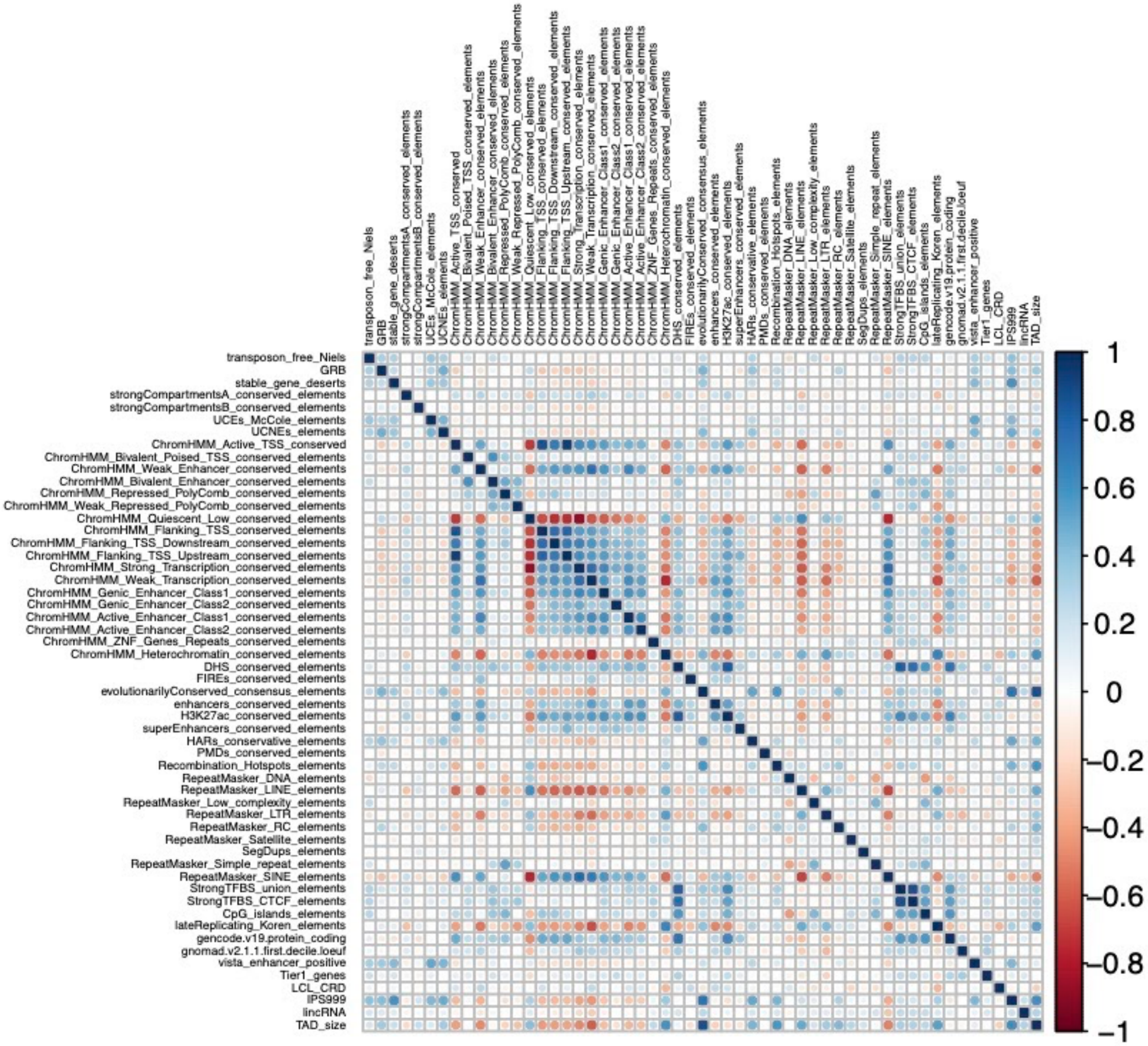


**Supplementary Figure 8. Correlation between genomic features tested for association with LRPEs.** Spearman correlation matrix for 54 features across five functional categories tested for an enrichment in TADs preferentially disrupted by case BCRs.

### METHODS

#### ***Cohort ascertainment and phenotyping***

We collected a cohort of 710 unrelated individuals with a BCR detected by karyotype from multiple sources, including through the International Breakpoint Mapping Consortium, the Danish Cytogenetic Central Registry, Developmental Genome Anatomy Project (DGAP), cases/controls previously reported in the literature by our group and others, as well as from the Coriell Institute for Medical Research (Supplementary Table 1). Affected individuals (referred to as "cases" throughout the manuscript) were defined as those ascertained for a DD and/or congenital anomaly (n=406) with a *de novo* or phenotype segregating BCR. Unaffected controls (n=304) were ascertained as a population control (n=143; 47.0%), an individual with idiopathic infertility (n=122; 40.2%), or as an unaffected parent transmitting an unbalanced BCR to a child (n=39; 12.8%). There were 49 families (n=15 cases and n=34 unaffected controls) where multiple individuals from both sexes had inherited the same inversion (only one individual from each family was included in the analyses). For the remaining 391 cases, 47.6% (n=186) were female and 52.4% (n=205) were male. Of the 270 remaining controls, 56.7% (n=153) were female and 43.3% (n=117) were male (Supplementary Table 1). We excluded all males with oligo/azoospermia and females with premature ovarian failure from our cohort given that they did not meet criteria for either case or control status. All studies were approved by respective institutional review boards (IRB). Informed consent was obtained from all subjects or their legal representative for participation in the study when the IRB so required.

#### ***BCR breakpoint mapping and annotation***

The vast majority of the cases and controls included in this study were mapped using liWGS.[5] The subset of cases and controls that were ascertained by DGAP were analyzed following a previously described protocol[6,7] and computational pipeline.[8,9] While the library preparation protocols differed slightly between sites and studies, the resolution (1-7kb) of each method was consistent. For the purposes of this study, we only included individuals with a simple BCR and removed all individuals with a complex rearrangement involving three or more breakpoints, a large copy number variation at the breakpoint, or individuals with a common founder inv(10).[10] For DD cases with Sanger sequencing available, 90.2% (148/164) of the BCR breakpoints resolved to less than 5kb of genomic imbalance at the breakpoint. All breakpoint coordinates are provided using the Genome Reference Consortium February 2009 build of the human genome (GRCh37/hg19) and annotated for gene disruptions using Gencode v19 annotations[11] based on the overlap between a breakpoint and any gene body (exon, intron, or UTR) as determined by BEDTools.[12] We further classified all protein-coding genes in the genome into four tiers based on their association with disease (described in Supplementary Table 3).[12]

#### ***BCR simulations***

We generated a simulated set of BCRs to model null expectations if BCRs were randomly distributed throughout the genome. To generate a set of simulated BCRs, we randomly sampled 30,400 pairs of coordinates from all alignable sequences in the primary assembly of the reference genome while matching on the properties of the 304 BCRs empirically identified in control individuals, as described above. Specifically, we matched on the proportion of translocations versus inversions, and further matched on inversion size (*i.e.*, the distance between breakpoints). When sampling random coordinates from the genome, we excluded any region present in the ENCODE short-read mapping exclusion list,[13] any N-masked reference sequences, or any region with <50% alignability by 100mer reads per the UCSC Browser.[14] In total, we excluded 394Mb (or 12.7%) of the total GRCh37 reference. Over 95.5% of the empirical BCRs did not overlap any of these excluded regions.

***<u>Genome-wide BCR breakpoint analyses</u>***
We conducted several analyses to evaluate the distributions of BCR breakpoints throughout the genome, grouping BCRs per chromosome, per chromosome arm, and per 862 cytobands taken from the UCSC Genome Browser.[15] For these analyses, we compared subsets of breakpoints (*e.g.*, cases vs. controls, or cases vs. simulated BCRs) using two-tailed Fisher's exact tests and assessed significance at a Bonferroni-adjusted threshold accounting for all tests. Separately, we also conducted analyses normalized for chromosome length. We normalized each BCR breakpoint coordinate relative to its chromosomal position by first computing the distance between the breakpoint and that chromosome's centromere using centromere coordinates from the UCSC Genome Browser before dividing that distance by the length of the corresponding chromosome arm. In "signed" analyses, we assigned negative signs to all coordinates localized to chromosomal p-arms whereas breakpoints localized to q-arms were left as positive values. Finally, we compared distributions of chromosome length-normalized BCR breakpoints between subsets (*e.g.*, cases vs. controls) using a two-sided Kolmogorov-Smirnov test and correcting *P*-values for multiple comparisons.

***<u>Gene-based burden tests</u>***
We compared rates of BCRs disrupting several different gene sets (*e.g.*, gene Tiers 1-4, described above) in cases, controls, and simulated BCR carriers using two-sided Fisher's exact tests of counts of BCRs that did or did not disrupt at least one gene per gene set and assessed significance at a Bonferroni-corrected threshold that accounted for all gene sets tested. We also evaluated evidence of association for each protein-coding gene in cases vs. controls and cases vs. simulated BCR carriers with a similar approach: we conducted two-sided Fisher's exact tests of counts of BCRs disrupting each gene in cases versus either controls or simulated BCR carriers and assessed all enrichments at exome-wide significance accounting for all genes tested.

***<u>Identification of noncoding mechanisms underlying pathogenic LRPEs</u>***
We first assessed whether BCRs from DD cases were more likely to disrupt noncoding genes than controls by annotating all BCR breakpoints for overlap with Gencode v19 gene annotations[42] and comparing the fraction of cases versus controls disrupting various classes of noncoding genes (all genes, just lincRNAs, *etc.*) using Fisher's exact tests. Second, we investigated whether proximity to a known disease gene was sufficient to discriminate case versus control status by comparing the proportion of cases versus controls with a breakpoint near genes in each gene tier using a two-sample Kolmogorov-Smirnov test. Given that most structural variants associated with LRPEs are within ~2Mb

of their target gene, we conservatively excluded cases and controls with a BCR breakpoint at a distance >5Mb of a gene in each gene tier in the analysis. Next, to see if 3D topology may be more informative than linear distance, we compared the proportion of cases versus controls with a breakpoint in a TAD harboring at least one gene from each of our gene tiers using a Chi-square test. Both analyses were performed with and without direct gene disruptions for each gene tier.

***Genome-wide association of TADs with DDs***

To determine if there are any individual TADs with an accumulation of BCR breakpoints beyond what would be expected by chance, we intersected our breakpoint atlas with 2,257 autosomal TAD boundaries identified from Hi-C in a fetal lung fibroblast cell line (IMR90).[2] Given the sparsity of cytogenetic BCR breakpoints across the genome, we remain under-powered to perform direct association tests between cases and controls and therefore calculated a *P* value for each TAD by comparing the observed number of cases with a breakpoint in that particular TAD to an expectation based on a null Poisson model derived from the distribution of control BCRs. Bins on the X and Y chromosomes were removed, and we corrected for TAD size and case-control cohort imbalance. We defined a Bonferroni-adjusted $P$ value $\leq 2.2 \times 10^{-5}$ by correcting for the total number of TADs tested. We also defined a second set of TADs with suggestive evidence for association with case status based on a BH-FDR < 10%. To confirm that the genome-wide significant loci were not driven by cell-type specific TAD boundaries identified from the IMR90 cell line or technical confounders of the underlying Hi-C data, we performed the analysis described above using TAD boundaries identified from five additional tissues as well as the densest Hi-C map generated to date, from the GM12878 lymphoblastoid cell line (Supplementary Fig. 5).[1,3,16]

***Generation of phased Hi-C maps from individuals with noncoding BCRs disrupting the TAD containing MEF2C***

We generated Hi-C libraries on five cases with noncoding BCRs disrupting the TAD containing *MEF2C* following previously published protocols.[3,17] To simultaneously assemble, phase, and identify SVs we developed a 3D resequencing workflow that is based on the 3D *de novo* assembly (3D-DNA) pipeline[18] and the Juicebox Assembly Tools[19] ecosystem. The workflow begins with identifying local variants such as SNPs and indels using the DRAGEN variant caller from Hi-C data.[20] Next, the local variants are assigned to homologs with the 3D-DNA phaser.[17,21] The phased variants are then used to create a Hi-C map illustrating contact patterns between DNA sequences in a haplotype-specific fashion. This haploid Hi-C contact map can then be examined in Juicebox Assembly Tools to identify, annotate and resolve large-scale rearrangements, in a molecule-specific fashion. This allows us to reconstruct end-to-end sequences for each chromosomal homolog that reflect both the local variation and the large-scale sequence rearrangement and build a Hi-C contact map reflecting the “true” contact pattern for each homolog, whether rearranged or not. Note that there is some ambiguity in the end-to-end sequences because a small subset of variants cannot be reliably phased.

***Identification of features associated with TADs intolerant to disruption***

The substantial fraction of controls with a BCR breakpoint directly disrupting a TAD harboring a Tier 1 gene suggests that other features beyond genic content are likely contributing to the risk for pathogenic LRPEs. To test this hypothesis, we compiled a list of 54 coding and noncoding features (Supplementary Table 9) to test for enrichment in TADs preferentially disrupted in cases compared to controls. To test as many hypotheses as possible, we included features from multiple functional categories, including genes, *cis*-regulatory elements, conserved elements, repetitive elements, and "other". To account for tissue-specificity, we only included elements that were identified in >50% of the 75 tissues used to define the 18 chromatin states from the Epigenomics Roadmap dataset.[22] We defined three possible types of overlap for the annotations, including count (the number of features overlapped by the TAD), overlap (the fraction of the TAD overlapped by the feature), and binary (presence/absence of the feature in the TAD). The type of overlap chosen for each annotation was determined after visualizing the distribution of the feature across all TADs in the genome. Any continuous variables that did not follow a normal distribution were $\log_{10}$ transformed before inclusion in the analyses.

To look for features potentially associated with TADs intolerant to disruption, we defined a set of training data consisting of TADs disrupted by two or more BCR cases and no BCR controls (positive training data; n=45) and TADs disrupted by one or more BCR control and no BCR cases (negative training data; n=261). Using this training data, we first performed a univariate logistic regression analysis using the stats R package (version 4.0.2) for each of the 54 annotations and considered any feature with a BH-FDR < 5% as being potentially associated with case status. Next, we performed an elastic net regression using the glmnet R package (version 4.0-2) with the training data as input to identify features that were predictive of case status after controlling for the effects of all other features being considered. We set the alpha value to 0.5 and performed cross-validation to determine the best lambda value for the elastic net regression.

***LRPE model validation***

We applied our six feature LRPE model to all 2,257 autosomal TADs in the genome and used two separate approaches to validate its predictive accuracy. First, we defined an independent set of positive and negative test TADs, which included TADs disrupted by one or more BCR cases and no controls (n=178; "positive" TADs). Given that we had used all of the control BCR breakpoints to define the negative training data, we leveraged a second SV data set, gnomAD-SV,[23] to define the negative test TADs. We considered any TAD that had one of its boundaries overlapped by a high-quality (PASS, QUAL>500, read-depth support), common (>1% frequency), deletion >10kb obtained from gnomAD SV as likely tolerating disruption (n=198, "negative" TADs) and compared the performance of the model using receiver operating characteristic and precision-recall curves. Second, we ranked each TAD in the genome based on our model and compared the cumulative case-control, case-simulation, and control-simulation enrichments across five-percentile bins. We defined a "LRPE pathogenicity" cutoff score based on the point in which case-control and case-simulation enrichments surpassed OR>1.5 (Fig 6D), which corresponded to TADs ranked in the top 10th percentile or higher. We searched the literature for known pathogenic LRPE regions that were not represented in our training data and identified eight loci (Supplementary Table 10) that were disrupted by two or more independent cases with noncoding BCRs. For each loci the target gene also had to be identified and molecularly confirmed (*e.g.*, dysregulation of the target gene observed in patient cell lines). We overlapped the known pathogenic LRPE loci with TAD boundaries and compared the fraction of these eight TADs that fell above the cutoff score compared to all other TADs in the genome using a Fisher's exact test.
